# Lipoprotein profile and metabolic fine-mapping of genetic lipid risk loci

**DOI:** 10.1101/2022.06.12.22276286

**Authors:** Karsten Suhre, Raghad Al-Ishaq, Aziz Belkadi, Tanwir Habib, Anna Halama, Nisha Stephan, Gaurav Thareja, Shaza Zaghlool, Eric B. Fauman, S. Hani Najafi-Shoushtari

**Affiliations:** Bioinformatics Core, Weill Cornell Medicine-Qatar, Education City, 24144 Doha, Qatar; Department of Biophysics and Physiology, Weill Cornell Medicine, New York, NY, U.S.A.; Internal Medicine Research Unit, Pfizer Worldwide Research, Development and Medical, Cambridge, MA, 02139, USA; Division of Research, Weill Cornell Medicine-Qatar, Education City, 24144 Doha, Qatar; Department of Cell and Developmental Biology, Weill Cornell Medicine, New York, NY, U.S.A.

**Keywords:** Lipid and cholesterol metabolism, Lipoprotein particle composition and remodeling, Cardiovascular disease, Genetic association, Drug target evaluation

## Abstract

Dysregulated blood lipid levels sit at the nexus of cardiometabolic disorders and are major predictors of human cardiovascular health. Using five major lipid traits (HDL-C, LDL-C, non-HDL-C, TC, and TG), a recent genome-wide association study (GWAS) in 1.65 million individuals identified and fine-mapped over 1,000 genetic loci that may be implicated in the etiology of dyslipidemia and related cardiovascular disease. However, a deeper functional understanding of these associations is needed to assess their therapeutic potential as druggable targets. Here we leveraged data from over 98,000 participants of UK Biobank for deep molecular phenotypic refinement and identified 225 lipid risk variants that associated with 168 distinct NMR-derived lipoprotein and metabolic traits, doubling the number of loci that are discoverable when using the five “classical” lipid traits alone. Hypothesis-free testing of >14,000 ratios between metabolite pairs significantly increased statistical power (p-gain) at 72% of the loci, revealing distinct groups of variants with functionally matching NMR-ratios that affect lipoprotein metabolism, transport, and remodeling (LPmtr). We demonstrate how these NMR- trait and -ratio associations can be used in the functional interpretation of the respective lipid risk loci and their evaluation as potential drug targets. Our study reveals a comprehensive picture of the biological roles that the different genetic variants play in LPmtr and supports the emerging view that lipoprotein size and core composition are essential for the understanding, prevention and treatment of lipid-related disorders, beyond the “classical” five major lipid traits currently used in clinical practice.

## INTRODUCTION

Improper trafficking of plasma cholesterol and other lipids in lipoprotein particles lies at the heart of metabolic dysregulation and is associated with atherosclerotic cardiovascular diseases (ASCVD) ^1^. Patients suffering from ASCVD often present with elevated LDL-cholesterol (LDL-and triglyceride-rich lipoproteins (TRLs) levels accompanied by reduced concentrations of HDL-Cholesterol (HDL-C) ^2–5^. While LDL-C levels are efficiently controlled by statins and the recently developed PCSK9 inhibitors, newer reports indicate that a large burden of ASCVD remains, due mainly to a lack of targeted therapies to alleviate persistent dyslipidemia caused by impaired hepatic clearance of TRLs (aka cholesterol remnants) from the arteries, and reduced cholesterol efflux capacity from foam cells, two major pro-atherogenic events that directly contribute to vascular inflammation and atherosclerotic plaque formation ^6–11^.

Moreover, while LDL-C levels are recognized as a major causal risk factor for ASCVD, Mendelian randomization studies ruled out a causal effect of clinical HDL-C levels on the risk of ASCVD, suggesting that HDL particles exert their anti-atherogenic activities and function through specific lipids and protein components other than their core cholesterol content ^12, 13^. Indeed, emerging evidence suggests that variations in HDL particle size and composition of lipids, proteins, and small non-coding RNAs affect HDL function under healthy and diseased conditions, underscoring the importance of identifying gene targets that contribute to the cardioprotective function of HDL subclasses ^14, 15^.

Over the last decade, genome-wide association studies (GWAS) have shed much light on common genetic variants associated with major lipid traits that were found to be implicated in key regulatory pathways underlying cholesterol and triglyceride metabolism, thereby identifying new modes of metabolic control in the pathogenesis of ASCVD, highlighting potential therapeutic targets in the reverse cholesterol transport pathway ^16^. Most recently, Graham *et al*. ^17^ published the largest GWAS with lipid levels in a multi-ethnic study with 1.65 million individuals, including 350,000 participants of non-European ancestries. They reported 1,765 index variants in 773 genomic regions that were associated with the five “classical” blood lipid traits, LDL-C, HDL-C, triglycerides (TG), total cholesterol (TC), and non-HDL-C ^17^. While the genetic architecture of these lipid-risk loci was mapped out in detail in their study, much less is known about their mechanisms, the metabolic pleiotropy of these loci, their potential link to ASCVD risk, and their role in lipoprotein metabolism, transport, and remodeling.

Here we leverage the recently published Nightingale NMR data that is now available for ∼100,00 participants of the UK Biobank to refine the metabolic architecture of these genetic lipid risk loci with a comprehensive set of 168 metabolite and lipoprotein related traits (Methods and **Supplementary Tables 1&2**) ^18–21^. We hypothesize that this kind of deep molecular phenotyping will shed new light onto the biological functions of the Graham *et al*. lipid risk genes and their role in regulating lipoprotein composition and turnover.

Our analysis includes three steps of genetic association analyses. First, we attempt a replication of the Graham *et al*. ^17^ lipid GWAS using the same five lipid traits as used in the original study but measured on the Nightingale platform. Then, we proceed to a molecular fine mapping of the replicated loci using the 168 NMR traits as endpoints. Finally, we conduct a hypothesis-free all-against-all ratio-metric association study, testing over 14,000 ratios at each of the Graham *et al*. genetic lipid risk loci.

To gain biologically relevant insights from these associations we deploy a formal fine-mapping approach to all loci and create credible sets of NMR traits that are most directly influenced by the associated genetic variants. We then use statin medication and incident myocardial infarction (MI) as a showcase to demonstrate how genetic NMR-trait association profiles can be used to predict the intervention outcome of presently less-well studied risk variants as genetic proxies for potential drugs. Finally, we focus our investigation on those genes that are directly involved in lipoprotein metabolism, transport, and remodeling (LPmtr genes) and identify NMR-ratios that reflect non-genetic variance that is shared by these LPmtr genes.

## RESULTS

### Metabolic fine mapping almost doubles the number of discoverable lipid risk loci

Graham *et al*. ^17^ reported 2,624 genetic associations on 1,835 lead variants with five major lipid traits (TC, TG, LDL-C, HDL-C, non-HDL-C). These associations included conditional analyses with up to 46 variants in the model and were conducted in multiple ancestries. Imputed genotype data for all but five of the variants were obtained from the UKB research analysis platform (RAP). We computed the associations of all 168 NMR metabolites and all 14,196 ratios between them in linear models with the lead variants, age, age^2^, sex, use of lipid lowering medication, the ten first genotype principal components, and all respective conditional variants reported by Graham *et al*. as covariates (**Supplementary Table 3**). While the conditioned models for the five lipid traits were sometimes different, their lead variants could be identical or in high LD. We therefore limited our analysis to lead variants that were not in strong LD (r^2^ < 0.7), and when multiple traits and conditional variants were available for the same lead variant, we kept the model with the strongest association signal in Graham *et al..* To avoid spurious associations with rare variants, we limited our analysis to minor allele frequencies of MAF>1% and further to associations that were genome-wide significant (p<10^-8^) in the conditioned analysis by Graham *et al*. In total, 1,054 lead variants satisfied these criteria constituted the starting point for the following analyses (**Figure 1**).

**Figure 1.**
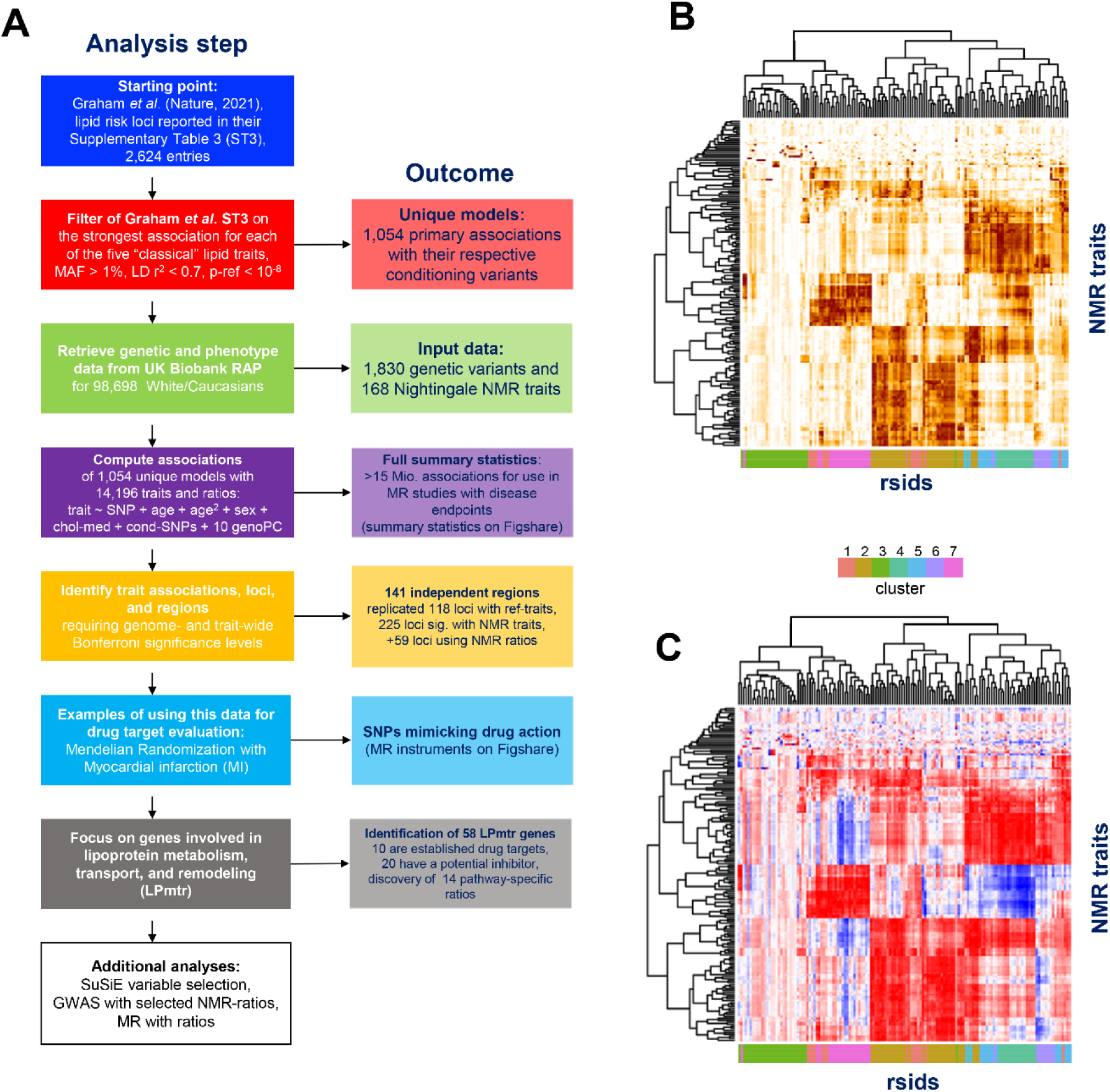
Study design and outcomes. (A) Flow chart of the present study; (B) Heatmap of negative log10(p-value) for the association of 168 NMR traits at the 141 independent genetic loci (rsids), scaled by division using the largest -log10(p-value) at each locus; Darker colors represent stronger associations; (C) Heatmap of the effect estimates (beta), scaled by division using the largest absolute beta value and aligning the directionality (sign of beta) such that the strongest beta at each locus has a positive association with the respective effect allele of that variant; Blue colors represent negative associations and red positive associations; Details on k-the means clusters are in **Supplementary Figure 4**; Heatmaps are also provided in Excel format as **Supplementary Tables 4&5**, see also **Supplementary Figure 3**.

Although we conducted association tests on a limited number of loci, we nevertheless chose to apply a genome-wide significance threshold of p_Bonf_ = 5×10^-8^ so that our conclusions also hold if conducted in a broader GWAS setting. We required conservative Bonferroni significance throughout by accounting for the number of traits tested as appropriate, that is, by dividing p_Bonf_ by 5 (p_ref_) for testing five lipids, by 168 (p_NMR_) for testing all NMR traits, and by 14,196 (p_AllRatios_) for testing all possible NMR ratios. To avoid rounding small p-values to zero, we report negative log10-scaled p-values throughout the paper.

A total of 118 out of the 1,054 primary associations were significant at p_ref_ for at least one of the five lipid traits reported by Graham *et al.* and can therefore be considered replicated – and even discoverable in a GWAS setting – in UKB using the Nightingale platform. The fact that we replicate only a fraction of the hits from Graham *et al.* is attributable to the reduced power of starting from a cohort that is sixteen-times smaller, and possibly to other population-specific factors, but not to the use of the NMR platform, as the NMR and the clinical biochemistry platforms yield highly correlated results (**Supplementary Figures 1&2**).

Of the 936 variants that did not replicate with any of these five “classical” lipid traits, 107 were significant at p_NMR_ with at least one of the 168 Nightingale traits. In other words, 225 of the 1,054 associations were significant at a genome-wide Bonferroni level with at least one of the 168 Nightingale traits. Hence, despite the increased multiple-testing burden, using the NMR traits led to the discovery of almost twice as many lead variants (225 versus 118) at loci that were established to be associated with lipid traits. One could argue that in these cases the respective associated NMR metabolite is then closer to the “true biology” of the implicate locus, and that the NMR association profiles contain a considerable amount of additional information compared to the five “classical” lipid traits that can be used to functionally fine map the Graham *et al*. lipid risk loci, as we show in the following.

### NMR association profiles reflect distinct biological pathways

Hypothesizing that genes at lipid risk loci with similar NMR association profiles share common biology, we decided to proceed with a k-means clustering of the -log10(p) association profiles. Based on gap statistics, an optimal number of seven clusters was chosen. We visualized the association statistics (-log_10_(p-value)) and effect sizes (aka betas) as clustered heatmaps with the lipid risk loci as one dimension and the Nightingale traits as the other (**Figure 1B/C** and **Figure 2**). To avoid counting similar signals at a single locus with several associated variants multiple times, we limited our analysis in the following to 141 sentinel variants that represent independent genetic regions, using a distance cut-off of 500kb to define regions and retaining only the variant with the strongest association signal to the NMR traits in each region. We found that many well studied genes that act on a same pathway clustered closely together.

**Figure 2.**
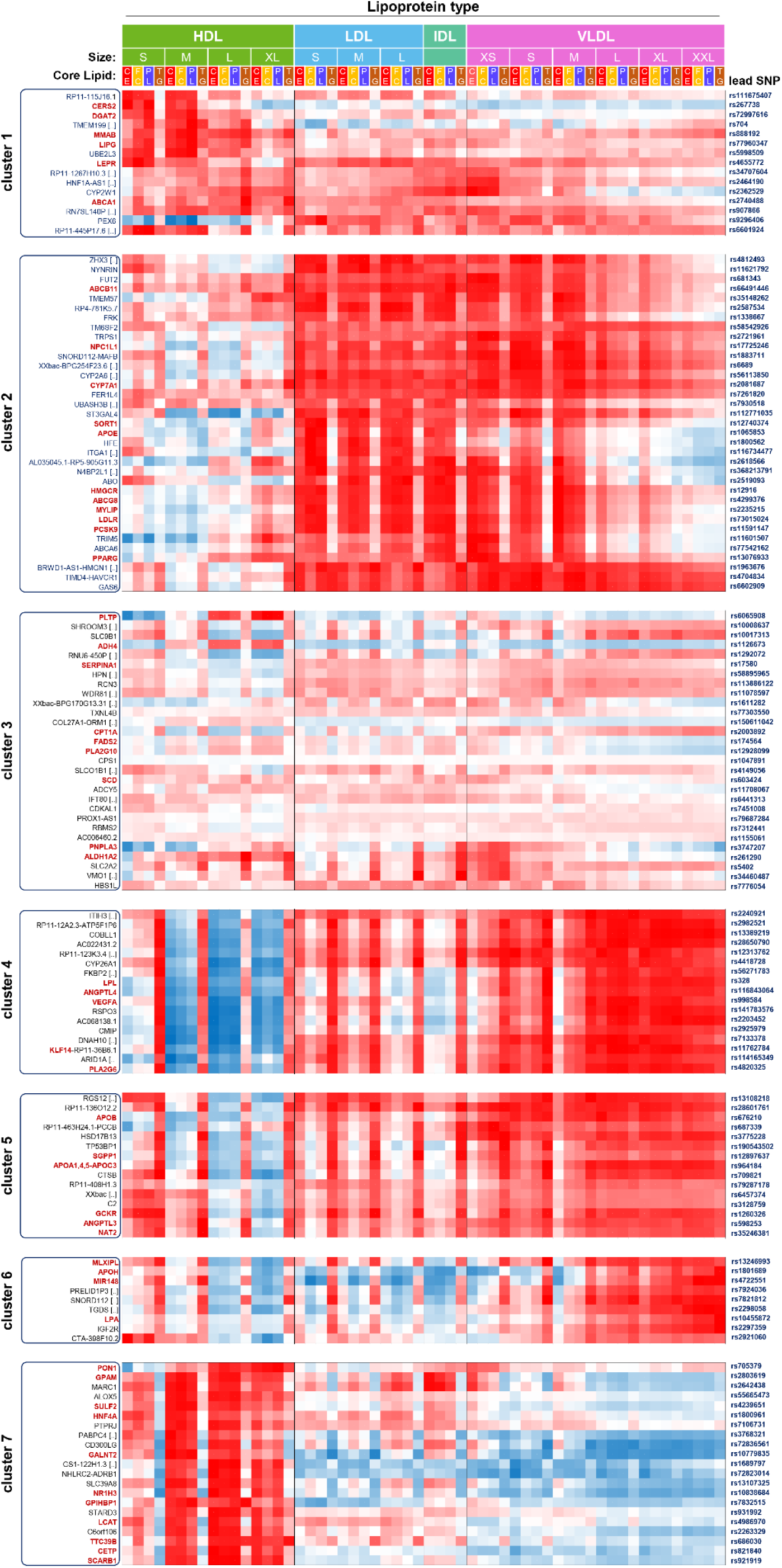
Association profiles of 141 independent genetic loci with the lipoprotein classes and LP core content. The association profiles (normalized effect sizes) are shown for cholesterol ester (CE), free cholesterol (FC), phospholipids (PL), and triglycerides (TG) for the fourteen lipoprotein size classes that are covered by the NMR platform, ordered by size from smallest (S-HDL) to largest (XXL-VLDL); Closest genes as identified by Graham *et al*. are in black; Genes annotated here as being involved in lipoprotein metabolism, transport, and remodeling are in red; Lead variants (rsids) are grouped by k-means cluster membership.

For instance, lecithin:cholesterol acyltransferase (LCAT) and cholesteryl ester transfer protein (CETP) are both enzymes critical to HDL maturation and remodeling ^22^. They share similar association profiles with the core composition of the 14 lipoprotein size classes and cluster closely together (cluster 7 in **Figure 2**). Both variants are predominantly associated with medium, large, and extra-large HDL particles, and to a lesser extent with the small HDL fraction, which is in accord with the established altered catabolism of apoA-1 and apoA-2 containing HDL particles in individuals lacking CETP ^23^ or LCAT ^24^. Both profiles showed a negative association with HDL-TG content and a positive association with HDL-PL, -FC and -CE, which is also in agreement with the established LCAT and CETP activity that ultimately results in FC and CE reduction and TG enrichment in HDL lipoprotein particles. Notably, cluster 7 also includes other HDL regulating factors, such as the sterol-responsive transcription factor LXR- alpha (*NR1H3*), *GALNT2* ^25^, *TTC39B* ^26^, and the HDL receptor *SCARB-1* (SR-B1) that controls the last step in HDL catabolism ^27^. *CETP* expression is activated by LXR-alpha, a central transcription factor that is destabilized and degraded by TT39B-mediated ubiquitination ^27^. The co-clustering of *CETP* and *TTC39B* suggests a potential biological link between these genes with possible ramifications for therapeutic T39 targeting and also suggests a role for the yet unknown co-clustered genes such as *SLC39A8*, *C6orf106* and *STARD3* in the regulation of HDL particle size.

Angiopoietin-like-4 (ANGPTL4), a secretory protein that restricts lipoprotein lipase (LPL) activity to limit plasma TRL-TG hydrolysis and subsequent release of fatty acids for uptake into adipose tissue during fasting, had a profile similar to LPL ^28^ (cluster 4, **Figure 2, Supplementary Figure 10C**). Loss of function variants and pharmacologic inactivation of ANGPTL4 is associated with reduced TG and increased HDL-C levels ^27, 29, 30^. Conversely, our data resembled a concordant inverse lipoprotein profile for the ANGPTL4 variant between HDL subclasses and their CE, PL and FC subtractions and TG, as reported for ANGPTL4 E40K loss of function mutation ^31^.

During feeding, ANGPTL3, by binding to ANGPTL8, acts in contrast to its ANGPTL4 counterpart, to replenish/restore fat depots in adipose tissue by blocking both endothelial lipase (LIPG) and LPL mediated TG-uptake through the liver and oxidative tissues ^32, 33^. Remarkably, we found that ANGPTL3 in cluster 5 is set apart from co-clustering with ANGPTL4 and LPL, despite sharing a similar LDL and VLDL profile (**Figure 2, Supplementary Figure 10F**). This is caused by differential correlation with small and medium HDL subclasses and is in accord with experimentally and clinically validated findings that show ANGPTL3 depletion or inhibition leads instead to a proportional reduction in HDL-C, non-HDL-C and TG levels ^30^.

Intriguingly, NMR associations revealed co-clustering of corroborated genetic variants associated with systemic and familial hypercholesterolemia that were not selectively captured in previous GWAS of human lipid traits. SNP rs73015024 near *LDLR*, a central factor of hepatic LDL-C and non-HDL-C uptake, SNP rs12916 near 3-hydroxy-3-methylglutaryl–coenzyme A reductase (HMGCR), the rate-limiting enzyme of cholesterol biosynthesis pathway and the target of LDL-C lowering statin drugs, and SNPs rs11591147 near *PCSK9* and rs2235215 near *MYLIP* (aka IDOL), two major factors facilitating post-translational LDLR degradation (cluster 2 in **Figure 2C**) ^34, 35^. Concordantly, we found that all four genetic variants were most strongly associated with all LDL subclasses and extra small to medium size VLDL particles resembling cholesterol remnants, confirming experimental findings that were not otherwise obtained from previous GWAS. The phenotypic correlations between selected pairs of these LDL loci (**Supplementary Figure 10ADE**) reveals rs2235215 as a novel gain-of-function mutation associated with *MYLIP* with potential therapeutic implications.

Data summarized in pre-formatted **Supplementary Tables 4&5** (see also **Supplementary Figures 3&4**) provide an analytical framework for the identification and fine-mapping of other associated gene(s) with NMR-measured lipoprotein-associated metabolic traits.

### Credible sets of NMR traits reveal the most direct associations at the lipid risk loci

The lipoprotein particle traits measured on the NMR platform, like for instance the concentration of esterified cholesterol in small VLDL particles, are determined by multiple processes, including cholesterol metabolism, fatty acid turn-over, phospholipid and triglyceride side-chain remodeling, and lipoprotein particle recycling, which can lead to intricated correlation structures between the traits. To account for possible bias in the clustering that may stem from these correlations, and to select NMR traits that are most representative for the observed associations, we applied the “Sum of single effects model” (SuSiE) approach ^36^. SuSiE is a Bayesian stepwise variable selection technique that is routinely applied in genetic fine mapping. Rather than picking the single most strongly independently associating variables, SuSiE identifies what are termed “credible sets” (CS) of variables (see methods). Applied to the NMR data, SuSiE identifies sets of traits that together share the maximal possible amount of non-redundant variance that can be explained by the respective genotype, or otherwise stated, SuSiE identifies those NMR traits that are likely to be the most directly affected by processes that are modulated by the variant (**Supplementary Table 6**).

The average number of credible sets identified for the 141 loci was 5.1 (median 3, maximum 24). We created a matrix with binary entries (0/1) that reflects which trait is a member of a CS at any given locus (**Supplementary Table 7&8** and **Supplementary Figures 5&6**). Unsurprisingly we found less correlations between the individual loci than when using the NMR association summary statistics for clustering. Albumin was the trait that appeared most frequently in the CS (32 times), followed by glycoprotein acetyls (26). It is interesting to note that these frequently associated traits were not generally among the strongest associations at the individual loci, but rather reflect the broader effect of variance in lipid metabolism on markers that correspond to processes such as free fatty acid transport in blood (albumin) and inflammation (glycoprotein acetyls). These CS can now be used to prioritize NMR traits to interpret specific lipid risk associations in the presence of highly correlated features.

### NMR association profiles can predict the therapeutic outcome of a medical intervention

A key application of GWAS in pharmaceutical drug development is the generation of genetic evidence to predict the efficacy of therapeutic interventions in humans. Extending the phenotype space from five lipids to 168 NMR traits is expected to represent a substantial gain in information. Each of the 225 NMR association profiles we generated here reflects a potential outcome of the therapeutic modulation of the underlying gene product.

As proof of concept, we evaluated the *HMGCR* locus. *HMGCR* encodes HMG-CoA reductase, the first committed step in cholesterol biosynthesis and the target of the statin class of lipid lowering agents. Statins are approved to reduce the risk of atherosclerotic cardiovascular disease and myocardial infarction (MI). To genetically evaluate the appropriateness of HMGCR inhibition to reduce the risk of MI we compared the association of each NMR species with incident MI to the impact of variants at *HMGCR* on those NMR species (**Figure 3**).

**Figure 3.**
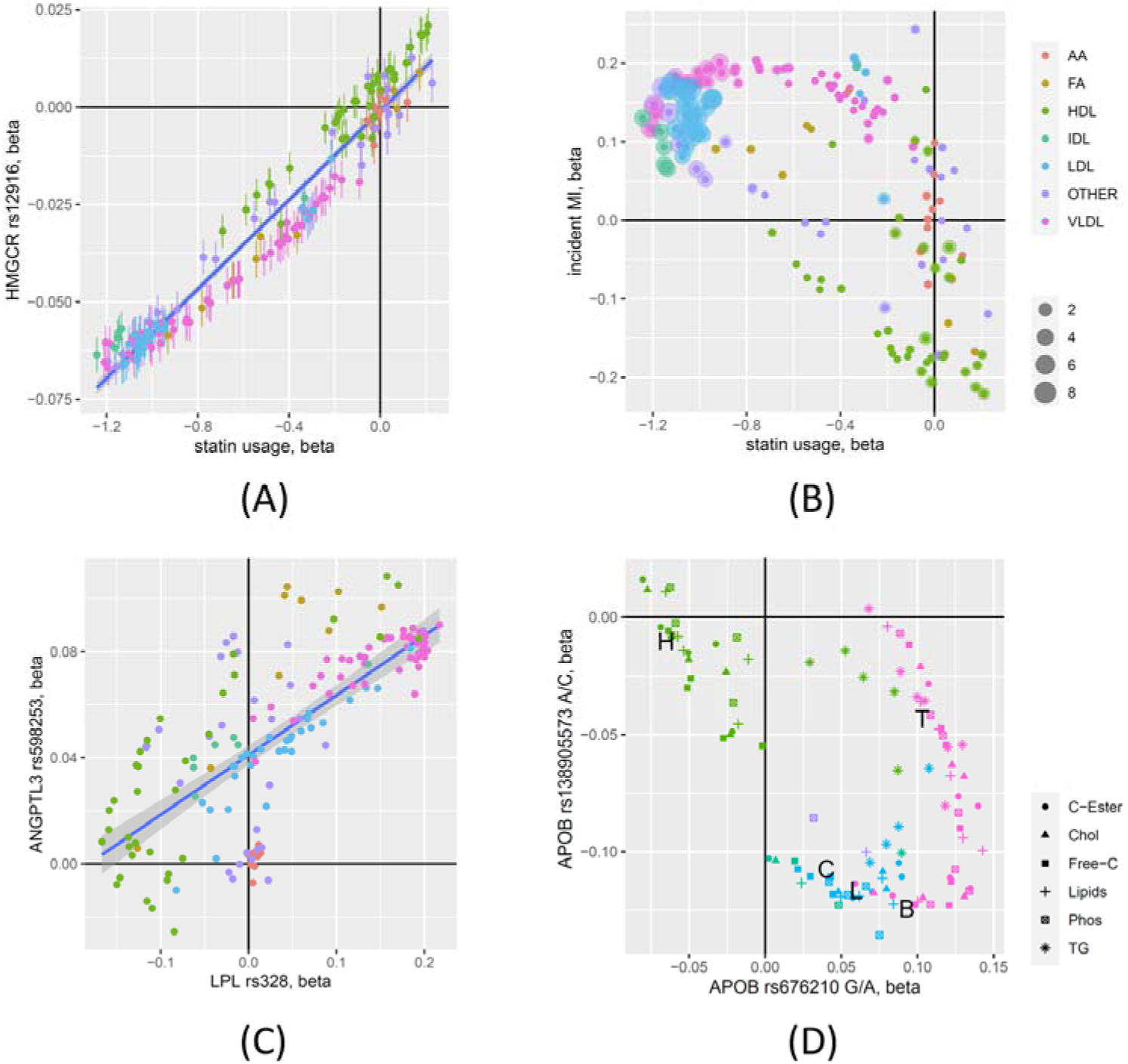
Application of NMR association profiles for drug target assessment. (A) Scatterplot of the effect estimates for the association of rs12916 near HMGCR with the 168 NMR traits and of the effect for the association of statin usage with the 168 NMR traits, colored by NMR trait group (**Supplementary Table 1**); Error bars represent the standard errors of the effect estimates; (B) Scatterplot of the effect size for the association of statin usage and incident MI with the 168 NMR traits; Dot sizes represent the log10(p-value) of the MR estimates (MR Egger) for the MR with MI as an outcome; **Supplementary Figure 7** shows the odds ratios for the associations with incident MI and estimated by MR; (C) Scatterplots of the effect sizes for the lead variants at the LPL and the ANGPTL3 loci with the 168 NMR-traits; (D) Scatterplot of the effect estimates for the association of the two independent variants (rs676210 and rs138905573) at the APOB locus with, limited to lipid traits (symbols) in the respective lipoprotein particles; The associations with the blood levels of HDL-C (H), LDL-C (L), total cholesterol (C), total triglycerides (T), and apolipoprotein B (B) measured by clinical biochemistry are indicated by the letters.

We first estimated causality for the NMR traits on incident MI. For this analysis, we excluded all individuals who reported any cholesterol-lowering medication at the time of recruitment. Of the 98,698 UKB participants for whom Nightingale data is available, 80,791 UKB participants did not use any cholesterol-lowering drugs. Among the UKB participants who did not use any cholesterol-lowering drugs at baseline, 1,100 subsequently had an MI event. To identify those NMR traits that are potentially causal for MI, we then conducted a Mendelian randomization study (MR) ^37^ with the NMR traits as exposures and MI as the outcome (**Supplementary Table 9** and **Supplementary Figure 7**). In total, 43 NMR traits had a significant (p<0.05/168) MR estimate using the conservative MR-Egger approach, 24 of which were related to LDL, eight to VLDL, five to IDL, and four to cholesterol traits. Apolipoprotein B (ApoB) was one of the strongest associations, in agreement with a previous study that prioritized ApoB as key lipid risk factor for coronary artery disease ^38^. Also consistent with previous studies ^13^, we found that no HDL-related traits were significant in the MR analysis. As a future application of our study, the NMR-trait exposure data we generated here can be used in MR analyses with different disease outcomes. These instruments are provided as **Supplementary Data** on FigShare at https://doi.org/10.6084/m9.figshare.19728991.

Next, we compared the impact of variants near *HMGCR* on the NMR traits identified as potentially causal for MI (**Figure 3A**). We found that in general, the traits most strongly influenced by *HMGCR* were those most strongly correlated with incident MI, especially the LDL-related traits. However, many VLDL-related traits had a stronger effect on MI risk but were less strongly influenced by *HMGCR*. The strongest risk factor not influenced by *HMGCR* variants is Glycoprotein Acetyls (GlycA). GlycA is an inflammatory biomarker that captures distinct sources of inflammation and provides a measure of cardiovascular risk ^39^.

We then compared the impact on the lipid traits obtained from lifelong genetic modulation of HMGCR with therapeutic intervention at the gene product of *HMGCR*. Of the 98,698 UKB participants for whom Nightingale data was available, 15,050 reported being on a statin treatment. We found that the impact on the lipid species due to statin usage was highly correlated with the effect of the genetic variant at *HMGCR* on that same lipid (**Figure 3B**), which is expected but reassuring, nonetheless. These results suggest that one can assess the causality of specific traits or biomarkers and identify variants and targets that favorably impact those causal traits which may translate into therapeutic benefits by targeting the gene products of the impacted genes.

We provide and discuss additional examples in **Figure 3C/D** and Supplementary Figures 8-11. Taken together, these examples demonstrate how NMR-profile analysis can be used to evaluate the potential outcomes of therapeutic modulation of the associated gene products and guide further investigations.

### Ratios between NMR traits reveal shared biology and uncover new loci

The hypothesis-free testing of all-against-all ratios between metabolite levels can increase the statistical power of metabolomics studies, well beyond what is lost by the increased level of multiple testing ^40^. When two metabolic traits share the same non-genetic information, taking their ratio will reduce the variance that a SNP could not explain. When the shared non-genetic information is large, this can lead to very large gains in power to detect a genetic association (quantified by the p-gain). Most interestingly, large p-gains reveal the presence of shared biology or other shared confounders between the traits, that can then be used to group genetic variants based on such – in general yet to be identified – shared biology or confounder.

Ratios between metabolic traits as independent variables in statistical hypothesis tests have been used for a long time. However, they are often considered mere normalization techniques, such as dividing metabolite levels in urine by creatinine to account for variability in urine concentrations or normalizing metabolite levels in cell extracts by “Bradford” total protein content to account for variability in metabolite extraction. Nevertheless, several GWAS with ratios between metabolite pairs identified associations with exceptionally large gains in the strength of their association that reflected biochemical conversions between the implicated metabolite pairs ^40–44^. Using this approach, we inferred the *FADS1* gene product as a desaturase of C20:3 to C20:4 fatty acids, while others found reduced enzyme activity of alkyl-DHAP in smokers ^45^, identified metabolic markers for drug screening ^46^, and unraveled genetic variance in blood-type related genes associated with fibrinogen protein phosphorylation ^43^. Interestingly, Nightingale already suggested 81 ratios between the directly measured traits as additional endpoints for analysis. We treat these ratios (termed NightRatios) separately as a benchmark, representing a targeted ratio-metric approach. There are hence to major motivations to analyze ratios: first, to increase power to detect genetic associations, and second, to identify groups of genes that share common non-genetic biological variance, that is, genes that can be hypothesized to act in a shared pathway.

We computed the associations of all 14,196 possible ratios between the 168 NMR traits with all lipid risk loci, using the p-gain statistic to identify significant ratios ^40^. The p-gain of an association of a metabolite ratio is defined as the smaller value of the two p-values for a pair of metabolite associations divided by the p-value of the association for their ratio. A p-gain of 10 has been shown to be the equivalent of an alpha level of significance of 0.05 for a single test ^40^. Out of the 225 variants that were associated with at least one NMR-trait, 162 variants had a Bonferroni significant p-gain at a genome-wide level (p-gain_AllRatios_ > 10 * 10^6^ * 14,196 = 10^11.2^), indicating that in over two thirds of the cases a ratio added significant information to the associations. Of the 829 associations that were not significantly associated with at least one of the NMR traits, 59 became significant at p_AllRatios_ with at least one ratio. On the other hand, eight loci that were significant at p_NMR_ did not reach the higher significance level required when using ratios. Hence, testing all possible ratios rather than only the NMR traits allowed us to identify 59 new loci, while eight loci with weaker signals were lost due to the increased multiple testing burden. Ratios therefore provided new knowledge at already known loci in 72% (162/225) of the cases, while at the same time increasing the power to discover entirely new loci by 23% (51/225).

### Fifty-eight lipid risk loci that are involved in lipoprotein metabolism, transport, and remodeling can be characterized by 14 associated pathways-specific ratios

Genetic evidence that supports potential mode of action is key in drug target prioritization ^47^. Using SNiPA ^48^, GeneCards ^49^, PhenoScanner ^50^, and general PubMed searches, we identified 58 gene loci among the 141 unique lead variants that encode proteins involved in lipoprotein metabolism, transport, and remodeling (LPmtr genes), and that are – given their functions, associated NMR traits, and absence of alternative candidates – most likely causal for the observed lipid trait associations. To keep this analysis focused on potential targets of lipid metabolism, we excluded here genes that were broadly related to other functions, like energy homeostasis, inflammation, and development. We used Pharos ^51^ to annotate the 58 LPmtr genes with NCBI gene summaries, UniProt function annotations, and drug target development status (**Supplementary Table 10**). Ten of these genes had the highest Pharos drug target development status (*Tclin*), that is, their gene products are already targeted by an approved drug (*ALOX5, NPC1L1, LEPR, ABCA1, HMGCR, PCSK9, PPARG, APOC3, VEGFA, ANGPTL3*). Twenty further genes had the second highest Pharos status (*Tchem*), indicating that at least one compound with an activity cutoff of < 30 nM targets their gene products (*ABCB11, ALDH1A2, APOB, CETP, CPT1A, CTSB, CYP26A1, CYP7A1, DGAT2, GALNT2, LCAT, LDLR, LIPG, LPL, NR1H3, PLA2G10, PLA2G6, SCARB1, SCD, SORT1*). Inhibitors of some of these proteins are currently in clinical trials ^52^. These 58 genes thus constitute high value drug targets.

We used NMR ratios to identify shared biological pathways between these genes with the expectations that these may ultimately provide a deeper understanding of their roles and interactions in lipoprotein metabolism, transport, and remodeling. We retrieved for each of the 58 loci the strongest associating ratio that reached Bonferroni significance (p-gain > 10^11.2^). We found 45 unique ratios. As some of these ratios were correlated, we clustered the resulting 58×45 matrix and retained 14 ratios that were consistently associated with all genes in a cluster block (**Supplementary Figure 12**). The ratio that associated with the largest number of genes was *Free Cholesterol in LDL / LDL Cholesterol*, that is, the percentage of free cholesterol in all LDL particles (13 genes), followed by the ratio *Concentration of HDL Particles / Total Concentration of Lipoprotein Particles*, that is, the percentage of HDL particles (9 genes), and the ratio of *Triglycerides in Small HDL / Triglycerides in Medium HDL*, that is, an indicator of relative TG content between small and medium sized HDL particles (6 genes). Except for three ratios that were associated with a single gene, all other ratios linked multiple genes together. We argue that these 14 ratios reflect biology shared by the respective associated genes and that the ratios are markers for the corresponding pathways.

To show that this is indeed the case, we annotated known interactions between the genes that associate with a same ratio and summarized their shared functions by a pathway annotation (**Figure 4** and **Box 1**). For instance, of the nine genes that are associated with the percentage of HDL particles, seven share the same transcriptional repertoire controlled by SREBPs and LXRs, and all are involved in mechanisms related to LDLR mediated LDL-C uptake and cholesterol transport. Noticeable is how the ratios are most often specific to a single set of genes, although there are of course also cases where a gene appears to act in more than one pathway. Taken together, we believe that this approach of utilizing NMR lipid trait ratios helps to systematically identify potential mechanistic links between specific gene variants and regulatory pathways that are involved in lipoprotein metabolism.

**Figure 4.**
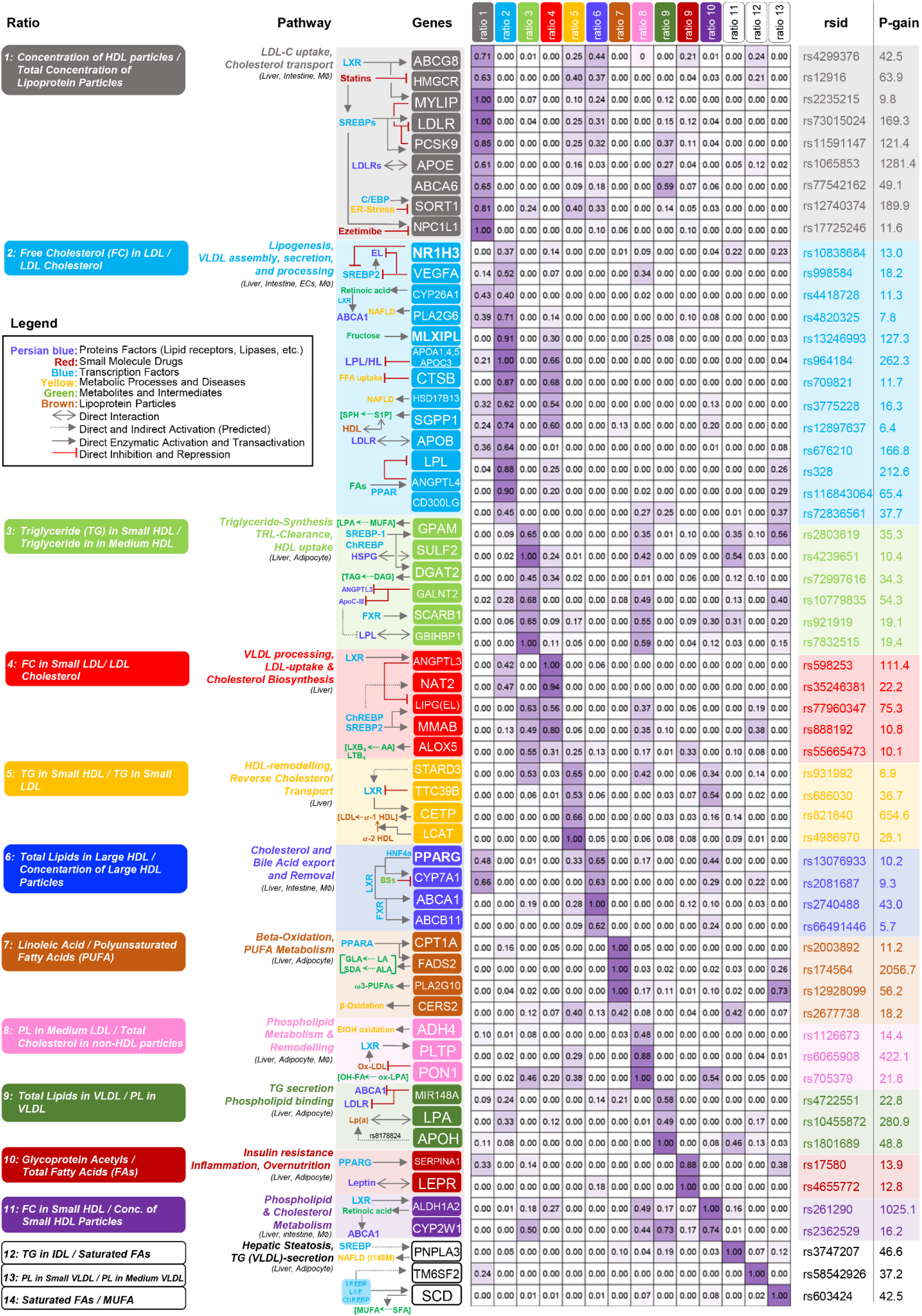
Genetic associations of 58 lipid metabolism, transport, and remodeling genes with 14 pathway specific ratios. Center: Heatmap displaying the relative log_10_(p-gain) for the associations of 14 key ratios with genetic variance at 58 lipid risk loci that harbor genes that are involved in lipoprotein metabolism, transport, and remodeling (LPmtr genes); Right: The highest log_10_(p-gain) observed at each given locus (rsid); Left: Full ratio names, matched to the 14 columns in the heatmap and functional annotation of the 58 LPmtr genes and pathways; refer to the embedded legend for symbols used in pathway annotations (detailed gene annotations and heatmap data are in **Supplementary Table 10**; see also **Supplementary Tables 11&12** for details on how this heatmap was obtained). Functional links between the ratios and gene groups are provided in **Box 1**.

#### Box 1. Biological insights into the pathways that link the 58 LPmtr genes and their link to the associated ratios.

Here we present selected highlights and bibliographic references that link the different LPmtr genes to their respective ratios. Abbreviations: reverse cholesterol transport (RCT), triglyceride-rich lipoproteins (TRL), free cholesterol (FC), cholesterol ester (CE), phospholipids (PL).

In this context it is interesting to note that the genes coding for the targets of three of the most successful cholesterol-lowering drugs (statins:*HMGCR*, ezetimibe:*NPC1L1*, and evolucumab:*PCSK9*) all associate with the same ratio (ratio 1 in Figure 4: the percentage of HDL particles). It might therefore be of interest to investigate still uncharacterized genes that associate with this same ratio for their potential role in lipoprotein metabolism, transport, and remodeling.

### Mendelian randomization with NMR ratios

We then asked whether NMR ratios might constitute causal markers for disease. We conducted a Mendelian randomization (MR) with MI as an outcome using the 144 unique lead NMR ratios associated at the 225 loci and that had a nominal p-gain > 10 as exposures (**Supplementary Table 13**). Overall, the estimated causal effect sizes and significance levels were weaker than those obtained using single NMR traits, the latter being dominated, as expected, by LDL traits. However, the third strongest MR association with ratios was the concentration of HDL particles divided by the total concentration of lipoprotein particles (p=3.2×10^-5^), whereas the single trait MR associations were substantially weaker (p= 0.008 and p=0.05, respectively). Interestingly, this is the ratio that we independently identified as ratio 1 in **Figure 4**. As the causal role of HDL-C in MI is unclear, we asked whether this ratio, which represents the percentage of HDL particles among all lipoprotein particles, was a predictor of incident MI in UKB. We found that statin-naïve UKB participants in the lower 10% range of HDL particle-% had twice the risk of having an MI in the follow-up period than the general UKB population (230 individuals compared to an expected 110 out of 1,100 statin-naïve UKB participants who had an MI after visiting the study center at baseline, chi^2^ p<10^-16^).

### A genome-wide association study with NMR ratios

Finally, we asked whether NMR ratios can reveal new associations. We therefore conducted a GWAS with the fourteen ratios reported in **Supplementary Table 10** and **Figure 4**. We identified 1,139 ratio associations with 595 genetically uncorrelated variants (**Supplementary Table 14**). For each of the 595 variants we then identified the most highly correlating variant in Graham *et al.* All but seven of the 595 variants were located on the same chromosome and within less than 10MB from a variant reported by Graham *et al*., indicating that using these ratios only very few new loci were discovered. However, over half of the variants that were located within 10MB of a known lipid risk locus (290 of 588) were not correlated (LD r2<0.1) with any of the Graham *et al*. lipid risk variants, suggesting that using these ratios additional genetic signals were discovered and that using ratios may be of value for further genetic fine mapping of these loci. This is in line with the expectation that, due to its much larger power, the Graham *et al*. study most likely already discovered all lipid risk loci that would be discoverable with ratios in our study.

An example of how ratios with large p-gains can generate new hypotheses regarding shared biology between the traits is *MLXIPL*, which encodes the carbohydrate-responsive element-binding protein (ChREBP), a transcription factor which binds to the carbohydrate response element (ChoRE) motifs in the promoters of triglyceride synthesis genes to instigate lipogenesis in response to high-carb diet ^53^. Matching its function, Graham *et al*. identified a strong association with TG at this locus that we also replicate here (rs3812316). New in this GWAS with ratios is the discovery of a strong association (p < 10^-118^) of the same variant with the ratio *Free Cholesterol in LDL / LDL Cholesterol*, that is, the percentage of free cholesterol in LDL particles. Note that the associations with the two individual NMR traits were barely significant (p>0.001). This observation suggests that genetic variation in *MLXIPL* not only leads to changes in TG levels, but also to a shift from free to esterified cholesterol in large LDL particles, which should be considered when investigating possible outcomes of medical MLXIPL inhibition.

## DISCUSSION

In this study we report the metabolic and lipoprotein fine-mapping of all currently known lipid risk loci using NMR measurements from the Nightingale platform, which is a dataset that has only recently been made available for over 98,000 samples from the UK Biobank. The benefit of having such detailed phenotypes for such a large number of samples becomes evident by the strength and breadth of the association signals that we report. Most striking were the many direct biological links that could be identified between the lipoprotein and metabolic traits and ratios and their associated gene variants that modulate many disease-relevant processes of lipoproteins metabolism, transport, and remodeling, as evidenced in **Figure 4** and **Box 1**.

A recent report of the European Atherosclerosis Society Consensus Panel concluded that the pathobiology of remnant lipoproteins is key to the development of optimal targeted therapies, but that understanding the atherogenic potential of their lipidomic and proteomic composition is very much a ‘work-in-progress’ ^1^. Our phenotypic fine mapping of the Graham et al. lipid risk variants responds to this challenge by generating biological insights at 225 independent gene loci. At over half of these loci the association strength increased by more than 9.5 orders of magnitude (p_NMR_) with an NMR trait compared to the five “classical” lipid traits, indicating that for each of these loci additional biochemical information has been generated, arguably getting closer to the true biology of the respective genes and loci. Moreover, in over two thirds of the cases, ratios between NMR traits further increased the strength of association at a genome-wide scale and beyond, revealing shared biology in many cases.

Our study has of course also its limitations – the biggest being the curse of the “too-many results”. We clearly could not discuss all associations to the degree that they deserve. We therefore share all associations in different formats and degrees of condensationas **Supplementary Data** on Figshare at https://doi.org/10.6084/m9.figshare.19728991 (see also **Supplementary Figure 13**). These data can (among others) be used as instruments in future Mendelian randomization studies and for the evaluation of specific gene products for their potential as drug targets.

Another limitation reflects the choices that had to be made regarding the scaling of the traits, the covariates used in the models, significance cut-offs etc. We tried to follow as closely as possible previous work (i.e. Graham *et al*.), and to be as conservative as possible (i.e. always using Bonferroni correction and not analyzing rare variants). However, some choices, like the number of clusters, distance measures etc. used in the NMR-association profile clustering are to a certain degree *ad hoc*. We therefore did not put too much weight on the interpretation of the exact cluster boundaries, but rather see these as indicating helper tools in the interpretation and comparison of the NMR association profiles.

We included drug treatments as covariates which may introduce bias ^54^. Approaches to correct for it have been demonstrated with the UKB ^55^ but are not very practical here due to the large number of traits. **Supplementary Figure 14** presents a scatterplot of the effect sizes (beta) for the associations with the respective lead NMR trait at the 141 loci, with and without including lipid lowering medication as a covariate, that shows that potential bias due to this effect is limited in our study.

The validity of the Nightingale platform is critical to our study. While the NMR platform has been used in many previous studies ^18, 56^, we additionally confirmed that results obtained using the NMR platform concord with those obtained using clinical biochemistry. We found that the Pearson correlation between the readouts from both platforms was larger than r^2^ = 0.75 (**Supplementary Figure 1**) and that the correlation between the effect sizes for the 141 lead associations was r^2^ = 0.974. Furthermore, the correlation between the effect sizes obtained using the ∼98,000 UKB samples and those reported by Graham et al. were r^2^ = 0.953 when using the NMR data and r^2^ = 0.974 when using the clinical biochemistry data (**Supplementary Figure 2**).

Metabolic fine mapping of genetic lipid risk loci to provide evidence to support or reject potential drug targets, with Mendelian randomization studies to be used to anticipate the results of randomized trials ^57, 58^, is not new. This approach using the Nightingale platform has been pioneered in a study with 8,330 individuals ^59^. Gordillo-Marañón et al. validated and prioritized 30 lipid-related therapeutic for CHD, including NPC1L1 and PCSK9 ^60^. Richardson et al. ^61^ provided strong evidence of an effect of drug-based genetic scores on coronary artery disease (CAD) for seven out of eight drug targets (HMGCR, PCKS9, NPC1L1, CETP, APOC3, ANGPTL4, LPL, but not ANGPTL3). Similar single-gene studies were also conducted on HMGCR ^62, 63^, PCSK9 ^63^, and CETP ^64^. We replicate most of these earlier associations, which validates our generalization to all currently known lipid risk loci.

Although the prevalence of ASCVD has globally declined, this trend is threatened by persistent dyslipidemia mainly caused by the obesity epidemic and the conjoined increase in Type 2 diabetes. New treatments are needed as recent studies indicate that many patients suffering from ASCVD fail to achieve optimal LDL-C reduction, while existing clinical therapies to increase HDL function and lower TRLs and cholesterol remnants are only modestly effective and remain elusive. Identification of genes that contribute to changes in lipid and protein constituents of lipoprotein subpopulations under physiologic and pathologic conditions is a challenge and an unmet need to develop novel precision therapeutics to improve ASCVD and MI outcomes. Here we established a clear role for 58 genes that play a potentially causal role in lipoprotein metabolism, transport, and remodeling. For the gene products of 30 of these genes an inhibitory molecule already exists that can now be further investigated. More generally, as we have shown in examples, our data can now be used to transfer knowledge from known to new lipid risk loci based on similarity of their respective NMR-trait and -ratio association profiles. Taken together, our deep molecular fine mapping of the Graham *et al*. lipid risk loci provides a comprehensive resource for the research into and the development of future lipid regulating drugs and treatment options.

## METHODS

### Data sources

All data was obtained through the UKB RAP system on the DNAnexus platform (data dispensed on December 14, 2021; application id 43418). Samples for analysis were selected using the following criteria: “Ethnic background | Instance 0” is “White”, “Genetic ethnic grouping” is “Caucasian”, “Spectrometer |Instance 0” is not NULL and “Measurement Quality Flag | Instance 0” is NULL, yielding NMR data for 98,698 out of 502,414 UKB participants. Imputed genotypes for 1,835 variants were extracted from UK RAP BGEN files (https://biobank.ndph.ox.ac.uk/showcase/label.cgi?id=100319) using bgenix ^65^ and reformatted to text format using plink ^66^. NMR data (https://biobank.ndph.ox.ac.uk/showcase/label.cgi?id=220) and additional phenotype data (age, sex, use of cholesterol lowering medication, medication usage, clinical biochemistry, …) were extracted using the DNAnexus cohort browser and the table downloader app (https://ukbiobank.dnanexus.com/landing). Incident myocardial infarction was defined as present when the reported “Date of myocardial infarction” (https://biobank.ndph.ox.ac.uk/showcase/field.cgi?id=42000) was later than the “Date of attending assessment centre” (https://biobank.ndph.ox.ac.uk/ukb/field.cgi?id=53). For all variables for which multiple instances were available, “Instance 0” (baseline) was selected.

### Lipoprotein and metabolic data

A detailed description of the metabolic traits and pathways covered by the Nightingale NMR platform is provided in **Supplementary Table 1** and https://biobank.ndph.ox.ac.uk/showcase/label.cgi?id=220. In brief, the platform readouts include several amino acids (alanine, glutamine, glycine, histidine, isoleucine, leucine, valine, phenylalanine, tyrosine), glycolysis related metabolites (glucose, lactate, pyruvate, citrate), ketone bodies (3-hydroxybutyrate, acetate, acetoacetate, acetone), creatinine, and two lipid species (linoleic acid, docosahexaenoic acid). The Nightingale platform further reports aggregated lipid traits, specifically, total fatty acids and their degree of unsaturation, omega-3 and omega-6 fatty acids, polyunsaturated fatty acids (PUFA), monounsaturated fatty acids (MUFAs), saturated fatty acids, phosphoglycerates, total cholines, phosphatidylcholines (PCs), and sphingomyelins (SMs). Moreover, the platform also provides readouts of protein glycosylation (glycoprotein acetyls), apolipoproteins B (ApoB) and A1 (ApoA1), and albumin. However, the largest part of the platform’s traits are quantitative measures of total lipids, free and total cholesterol (FC, TC), cholesteryl esters (CE), phospholipids (PL), and triglycerides (TGs), all segregated by lipid particle size, together with the concentrations of these particles. Lipoprotein particle classes cover fourteen size ranges, from small HDL particles with an average diameter of 8.7 nm to chylomicrons and extremely large VLDL, with particle diameters from 75 nm upwards. In addition to the individual traits, Nightingale suggests the computation of 81 ratios between these directly measured traits as additional endpoints, including the ratios of TGs to phosphoglycerides, apoB to apoA1, PUFAs to MUFAs, omega-6 fatty acids to omega-3 fatty acids, and the percentages of different lipid content in the lipoprotein size classes, bringing the total number of features up to 249 (**Supplementary Table 2**).

### Statistical analysis

The NMR data was log-scaled and ratios were computed using the identity log(A/B) = log(A) - log(B), which renders the association statistics invariant to inversion of nominator and denominator. NMR values equal to zero were treated as missing. The fraction of missing and zero values was very low, with 16,180 NA’s (0.098%) and 4,836 zero values (0.029%) out of 98,698*168 = 16,581,264 individual datapoints. The data was inverse-normal scaled after log-scaling to avoid overly strong effects of outliers that may have resulted from division by small values. Linear models were computed using R (version 4.1.0) using the R-package maplet (version 1.1.1) ^67^, with the NMR data as dependent variables and all genetic variants reported in Supplementary Table 3 by Graham *et al.*, plus age, age^2^, sex, and use of lipid lowering drugs to the model (binary) as independent variables. For ratios between two NMR traits, the p-gain was computed as the smaller of the two p-values for the individual trait associations divided by the p-value for the ratio ^40^. Assuming a genome-wide multiple testing level of ∼10^6^ tests and requiring nominal significance (p<0.05, p-gain>10 in case of ratios ^40^), the following Bonferroni levels of significance were applied, depending on the context:

- p_ref_ = 5×10^-8^/5 = 10^-8^ for testing the five lipid traits (also used by Graham *et al.*),
- p_NMR_ = 5×10^-8^ / 168 = 10^-9.5^ for testing the NMR traits,
- p_NightRatios_ = 5×10^-8^ / 249 = 10^-9.7^ for testing NightRatios,
- p_AllRatios_ = 5×10^-8^ / (168*169/2) = 10^-11.5^ for testing all ratios & traits,
- p-gain_AllRatios_ = 10^6^ * 10 * (168*169/2) = 10^11.2^ for testing all ratios & traits. K-means clustering was performed using the R package factoextra (version 1.0.7).

### SuSiE credible set computation

The R-version of the ‘sum of single effects’ (SuSiE) model ^36^ was used (susieR, version 0.12.19) to conduct variable selection and to identify credible sets (CS) of variables. SuSiE is a Bayesian stepwise variable selection method that is widely used in genetic fine mapping. Rather than picking the single most strongly independently associating variable(s), SuSiE identifies what are termed “credible sets” (CS) of variables. In cases where there are multiple traits in a credible set, SuSiE provides a posterior probability for each individual trait to be included in the model (PIP values). The sum over the PIP values in each credible set sums up to one. For each genetic locus, the genotype (coded as numerical value 0, 1, 2) of the lead variant was used as the dependent variable (y) in the SuSiE calculation. The 168 NMR traits were used as independent variables (X) after the following pre-processing: Zero and missing values were imputed to the minimal occurring non-zero value. The NMR traits were then log-scaled, residuals were calculated in a model that comprised the variables age, age2, sex, use of lipid lowering medication, and the first ten genotype PCs as covariates and were then inverse-normal scaled. The R function “susie” was used with default parameters, except for the allowed maximum number of non-zero effects in the regression model, which was set to 25 to assure that all possible effects were included. Note that the largest number of eventually identified non-zero effects in our study was 24. All credible sets together with the corresponding PIP values are provided as **Supplementary Table 6** and the corresponding plots are described in **Supplementary Figure 5**. A clustering of the binarized PIP value matrix is in **Supplementary Figure 6** and numeric values are in **Supplementary Table 7.**

### Phenome-wide association data

Buergel *et al*. trained a neural network to learn disease-specific metabolomic states from the 168 UKB NMR traits to predict individual multi-disease outcomes ^68^. The authors provide the ranked variable importance for the NMR traits to predict 24 conditions, including common metabolic, vascular, respiratory, musculoskeletal and neurological disorders and cancers (Supplementary Table 12 in Buergel *et al*.). In addition, they provide the correlations of all NMR traits with a comprehensive selection of 40 clinical predictors, including in-depth blood measurements (Supplementary Table 12 in Buergel *et al*.). We merged these data sets with our trait associations to provide additional information about the different functional roles that the NMR traits play in other diseases (**Supplementary Table 8**).

### Association with statin usage and incident MI

The number of non-statin users was 80,791 individuals, defined as individuals not using any cholesterol lowering drugs, the number of statin users was 15,050 individuals. Individuals using cholesterol lowering drugs but not statins were excluded. Covariates used in the model were age, age2, sex, HbA1c and glucose. Association with incident MI was limited to 80,791 samples not using any cholesterol lowering medication, leaving 79,691 controls and 1,100 incident MI cases (date of reported MI > date of visit of the UKB assessment center).

### Mendelian Randomization

MR was conducted using the R-interface to MR-Base with MI as an outcome and NMR traits or ratios as exposures ^37^. GWAS summary statistics from CARDIoGRAMplusC4D ^69^ were accessed through MR-Base (trait ieu-a-798, 43,676 cases, 128,199 controls). We used the association summary statistics of 215 uncorrelated SNPs with 168 NMR traits to generate exposures. Only strong instruments (*ad hoc* p-value < 10^-10^) were retained. 157 of the 168 NMR traits had at least three strong instruments. The MR Egger method was used to compute MR estimates (**Supplementary Table 9** for traits & **Supplementary Table 13** for ratios). Summary statistics for the exposures are provided as **Supplementary Data** in MR-Base compatible format and can be used online version (http://app.mrbase.org) to conduct further MR analyses with other outcomes and to generate detailed reports for all traits.

### Genome-wide association study with selected ratios

We conducted a GWAS of the 14 key ratios that characterize the 58 LPmtr genes using the UKB RAP platform with the genotyped data set. We used generalized linear models implemented in plink (version 2) ^66^ with age, sex, lipid lowering medication use and the first ten genotype PCs as covariates, filtering variants on minor allele count > 100 and MAF > 1%. NMR traits and ratios between them were inverse normal scaled. The p-gain was computed as previously described ^40^. Associations with a p-value of p < 5×10^-8^ / 14 and additionally a p-gain > 10 were retained and clumped into independent genetic loci across the 14 ratios using an LD cut-off of r^2^<0.1, followed by clumping of correlated lead signals using an LD cut-off of r^2^>0.9, as previously described ^70^. Lead variants were annotated using PhenoScanner ^50^ and the corresponding highest correlating variant in ST3 in Graham *et al*. was identified using UKB RAP genotype data (**Supplementary Table 14**).

## AUTHOR STATEMENTS

### Data availability statement

All analyzed UK Biobank data was obtained through the UKB RAP system under application reference number 43418. All generated data are provided with the paper or are available online on FigShare (https://doi.org/10.6084/m9.figshare.19728991).

### Ethics statement

All data and samples were collected by UK Biobank protocols following all relevant guidelines and procedures and were shared with the UKB users under rules reviewed by the UK Biobank ethics committee and board.

### Code availability statement

Only publicly available software was used in the data analysis (R and Rstudio).

## Data Availability

All data produced in the present work are contained in the manuscript or available on FigShare at https://doi.org/10.6084/m9.figshare.19728991

## Acknowledgements

The authors wish to thank all UK Biobank participants for their contribution.

## Author contributions

Study design and data analysis: K.S.; interpretation of association data: R.A., A.B., T.H., A.H., N.S., G.T., S.Z., E.B.F.; manuscript writing: K.S. and S.H.N-Sh.; All authors contributed to the interpretation of results and critically reviewed the manuscript.

**Ratio 1:** The percentage of HDL particles (**ratio 1**) associated with key regulators of LDL-C uptake and cholesterol remnant clearance, including *LDLR* and its posttranslational regulators *MYLIP* and *PCSK9*. SORT1 promotes LDL-uptake in liver and macrophages by binding to LDL and directing its lysosomal degradation ^71, 72^, while restricting hepatic ApoB/VLDL secretion ^73, 74^. This ratio also associated with *NPC1L1* and *HMGCR*, two major established therapeutic targets for LDL-C lowering drugs. The inhibition of HMGCR by statins increases LDLR expression via SREBP transactivation, suggesting that **ratio 1** is driven by the more dynamic changes in VLDL and LDL plasma levels, which are mirrored by changes in the percentage of HDL particles. In total, 7 out of 9 genes of this group share the same transcriptional repertoire by SREBPs and LXRs. These transcription factors maintain cholesterol homeostasis in response to cellular sterol levels, indicating that this set of genes is responsible for mechanisms underlying LDLR mediated LDL-C uptake and cholesterol excretion. Genetic upregulation of these processes then appears to induce a decrease in the percentage of HDL particles. This group of genes contains three of the most successful cholesterol-lowering drug targets (statins:HMGCR, ezetimibe:NPC1L1, and evolucumab:PCSK9).

**Ratio 2:** Reduction in LDL-FC subfraction leads to the remodeling of LDL to smaller, more atherogenic particles, as they escape binding to LDLR and thus remain in circulation ^75–77^. We found that the fraction of FC in LDL-C (**ratio 2**) associated with genes that are involved in hepatic lipogenesis (*PLA2G6, HSD17B13, MLXIPL*), VLDL secretion and processing (*APOB, LPL, ANGPTL4, CTSB, VEGFA*), and cholesterol efflux (*CYP26A1*, *APOA1* locus ^78^, *SGPP1*). These genes are all mainly controlled by the LXR-alpha (NR1H3) transcription factor, which binds to retinoic acid receptor RXR and restricts SREBP-2 mediated cholesterol biosynthesis, while promoting VLDL secretion and processing as well as cholesterol efflux. Genetic variance in these processes can thereby explain the associated ratio, that is, the fraction of FC in LDL-C.

**Ratio 3:** The lipid acquisition of HDL particles occurs mainly after secretion. Thereby, the TG core content in HDLs is mainly derived from TRLs through concomitant action of lipases (e.g. LPL, associated with ratio 2) and CETP (associated with ratio 5). We found, however, that the TG content in small vs. medium HDL (**ratio 3**) associated with genes related to HDL binding to cell surface proteins (SCARB1, aka SR-B1), that selectively facilitate the internalization of its CE content (SULF2), that activate surface heparan sulfate proteoglycans (the LPL interaction partner GBIHBP1), and that activate PLTP and block the function of APOC3 and ANGPTL3 to activate LPL at postprandial state, when TG loading of HDL occurs (GALNT2) ^25, 79, 80^. In addition, our associations match the function of key TG-related enzymes (GPAM and DGAT2), emphasizing TG biosynthesis and secretion as upstream modulators of TG content in HDL particles.

**Ratio 4:** Identification of gene factors that alter the FC content in small atherogenic LDL particles are of therapeutic interest. We found a set of genes affecting the ratio of FC in small LDL vs LDL-C (**ratio 4**) that are primarily involved in regulatory pathways, such as the hydrolysis of VLDL/LDL TG content (*ANGPTL3, LIPG*) ^81^ and its LDLR mediated uptake (*MMAB, ALOX5*) ^82, 83^. Notably, ALOX5 was shown to be an activator of SR-B1-mediated HDL-C uptake ^83^. Indeed, we observe overlapping association of ALOX5 with ratio 3, namely TG in small vs medium HDL, affected by a set of variants including in the gene coding for SR-B1 (SCARB1). How ANGPTL3 impacts LDL-C levels remained unknown until more recently ^81^. Our data revealed a strong correlation between ANGPTL3 and the endothelial lipase (LIPG) and their joint association with **ratio 4**. Apart from its role in hepatic HDL catabolism through HDL-PL hydrolysis ^84^, our data confirms that LIPG plays a role in ApoB-containing lipoprotein metabolism, as has been indicated in refs ^85, 86^ and confirmed more recently in refs ^81, 87^.

**Ratio 5:** Recent studies suggest that HDL-C and TGs are the strongest determinants of cholesterol efflux capacity ^88^. In **ratio 5** we report on another cluster of genes implicated in TG metabolism, which is distinct from those found to affect ratio 3. Our data captures critical plasma HDL-C and reverse cholesterol transport (RCT) regulating factors, that is, LCAT, CETP, and TTC39B (T39), that alter the ratio between TG in small HDL and small LDL particles (**ratio 5**). While LCAT acts towards maturation of HDL particles, CETP potentiates CE/TG transfer in the HDL to LDL conversion, the ultimate last step in RCT ^89^.

**Ratio 6:** RCT is a key function of HDL and inversely correlated with LDL-C and ASCVD development ^90^. It comprises multiple steps and commences with cholesterol efflux from the periphery (e.g. macrophages) towards cholesterol uptake and fecal excretion as bile acids. The here associated cholesterol efflux pump, ABCA1, and the bile acid pump ABCB11B, constitute the rate limiting steps in the RCT pathway ^83^, thus most significantly control the overall lipid content in large HDL particles (**ratio 6**), which less anti-atherogenic as they contain both Apo-A1 and Apo-A2 ^79, 91^. Notably, larger HDLs have a stronger beneficial effect on RCT as compared to anti-atherogenic smaller HDLs, especially in a disease-specific context such as type 2 diabetes ^92^. Additionally, we found that PPARG, a transcription factor known to transactivate *ABCA1* and *ABCB11* expression via LXR ^93^ and FXR ^94^, respectively, as well as activating *CYP71A* expression, the central enzyme in the bile acid pathways that converts cholesterol to alpha-hydroxycholesterol, associated with **ratio 6**. Concordantly, anti-diabetic PPARG agonists such as thiazolidinediones and other glitazones have been shown to elevate the ApoA-2/1 ratio in concert with increased HDL-C levels, confirming the mechanistic link observed here between “larger” HDL and the genes associated with **ratio 6.** These observations suggest that the beneficial effect of PPAR agonists on lowering atherosclerosis progression in individuals with T2D ^95^ appears to be caused by improved RCT rather than anti-atherogenic small HDLs. Moreover, given PPAR gamma agonists’ detrimental side effects on weight gain and hepatic steatosis, therapeutic interventions that directly activate both *ABCA1* and *ABCB11* expression might prove more efficient to treat atherosclerosis.

**Ratio 7:** Linoleic acid (an omega-6 PUFA) is the substrate of FADS2. **Ratio 7** reflects its enzymatic function, in line with previous reports ^41^. During fasting, CPT1A facilitates entry of FAs into the mitochondria for oxidation, the transcription of which is stimulated by the PPARA-transcription factor ^96^, which is known to activate genes in the beta-oxidation pathway. Recent reports also implicate FADS2 as another direct target of PPARA. CERS2 catalyzes hepatic C16:0 ceramide synthesis that leads to impaired FA beta-oxidation in IR, NAFLD, and ASCVD. Conversely, PLA2G10 acts to release omega-3 PUFAs (EPA, DHA) from epithelial cells. These observations suggest that these genes (*CPT1A, FADS2, PLA2G10 and CERS2*) are all determining factors in the fraction of linoleic acid within all PUFAs (**ratio 7**).

**Ratio 8:** Further biological functions were confirmed at the example of **ratio 8** (PLTP, ADH4, PON1), that is, between PL in medium LDL and TC in non-HDL particles yet indicating unknown mechanisms linking PL metabolism with non-HDL lipoprotein surface PL content. The function of PLTP, that transfers PL from TRLs to HDL upon hydrolysis, is a key step in HDL maturation; however, its role in humans remains unclear.

**Ratio 9:** GWAS implicated miR-148a as the most highly conserved intergenic microRNA associated with human lipid traits. In liver, it inhibits both LDLR and ABCA1 to compromise cholesterol trafficking in and out of hepatocytes, thus contributing to abnormal LDL/HDL ratios ^97, 98^. The joint association with **ratio 9** suggests a potential link between *miR-148a*, *APOH* and *LPA* in the regulation of VLDL secretion, by modulating VLDL-PL content (**ratio 9**). A potential functional link between *APOH* and *LPA* is also supported by a recent study demonstrating that a genetic variant in *APOH* increases *LPA* expression ^99^.

**Ratio 10:** GlycA is an inflammatory biomarker and provides a measure of cardiovascular risk ^39^. Its link in **ratio 10** with total FA content to genetic variance in *LEPR* reflects the connection between the regulation of nutritional (fat) intake and obesity associated inflammation. Loss of *Serpina1* in mice changed inflammatory, lipid metabolism, and cholesterol metabolism-related genes in the liver ^100^.

**Ratio 11:** The two genes, *ALDH1A2* and *CYP2W1*, both associate with the content of FC in small HDL particles (**ratio 11**) and are both involved in retinoic acid metabolism. Intriguingly, several of the genes that associated with the fraction of FC in LDL-C (ratio 2) are regulated via the retinoic acid receptor RXR (see above).

**Ratios 12-14:** The ratio of *saturated fatty acids / monounsaturated fatty acids* corresponds to the substrate-product pair of SCD and reflects its enzymatic function ^43^ (**ratio 14**). *PNPLA3*, another SREBP target gene, most dominantly affects the ratio between TG in IDL and saturated fatty acids (**ratio 12**), while *TM6SF2*, a potential LXR target gene, appears to act towards the regulation of PL in small vs. medium VLDLs (**ratio 13**), pointing to a role in VLDL secretion and remodeling.

## SUPPLEMENTARY FIGURES

**Supplementary Figure 1:**
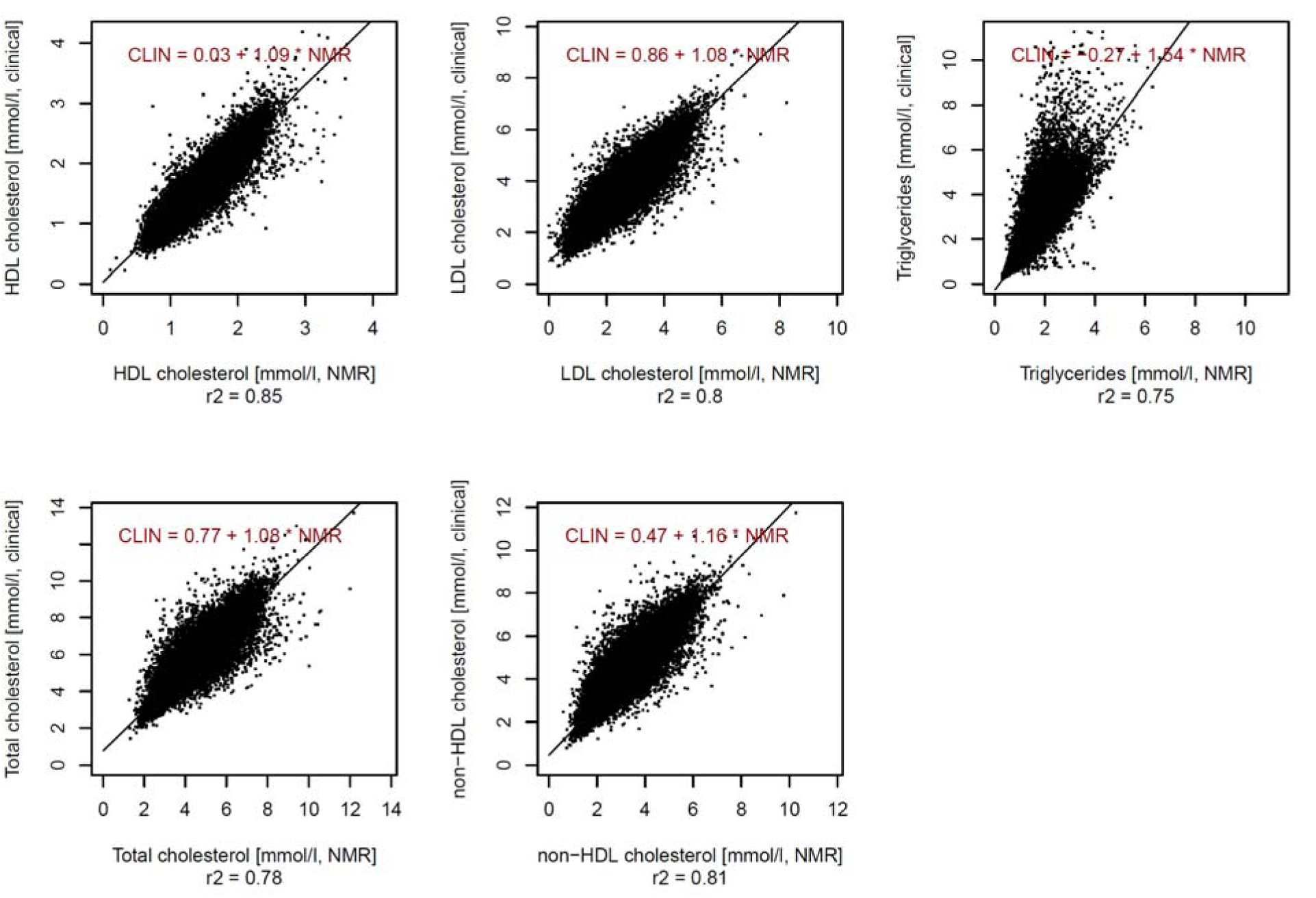
Scatterplots between the five lipid traits measured on the NMR platform and using clinical biochemistry methods show a high degree of correlation between both platforms (Pearson r2 > 0.75).

**Supplementary Figure 2:**
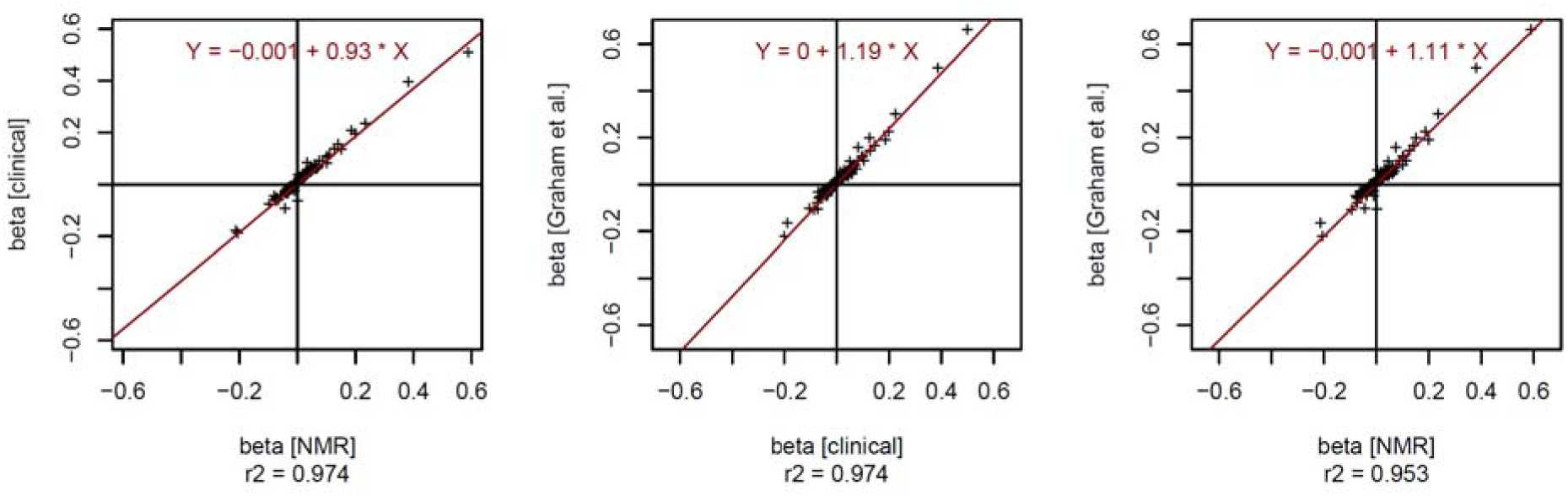
Scatterplots between the effect sizes (beta) for the associations of the 141 lead variants with the lead lipid trait reported in ST3 by Graham *et al*. obtained in different ways: Left: using the five lipid traits from the Nightingale NMR data (beta [NMR]) compared to using the clinical biochemistry data (beta [clinical]) on the 98,000 UKB samples; Middle: using the clinical biochemistry data compared to estimates from ST3 in Graham *et al*. (beta [Graham *et al*.]); Right: using the NMR data compared to Graham *et al*; These plots demonstrate that there is a very high correlation between the effect sizes (betas) derived using the NMR platform and the clinical biochemistry measurements on samples from 98,000 UKB participants, and that both also highly correlate with the effect sizes derived by Graham *et al*. using data from 1.6 million multi-ethnic individuals.

**Supplementary Figure 3:**
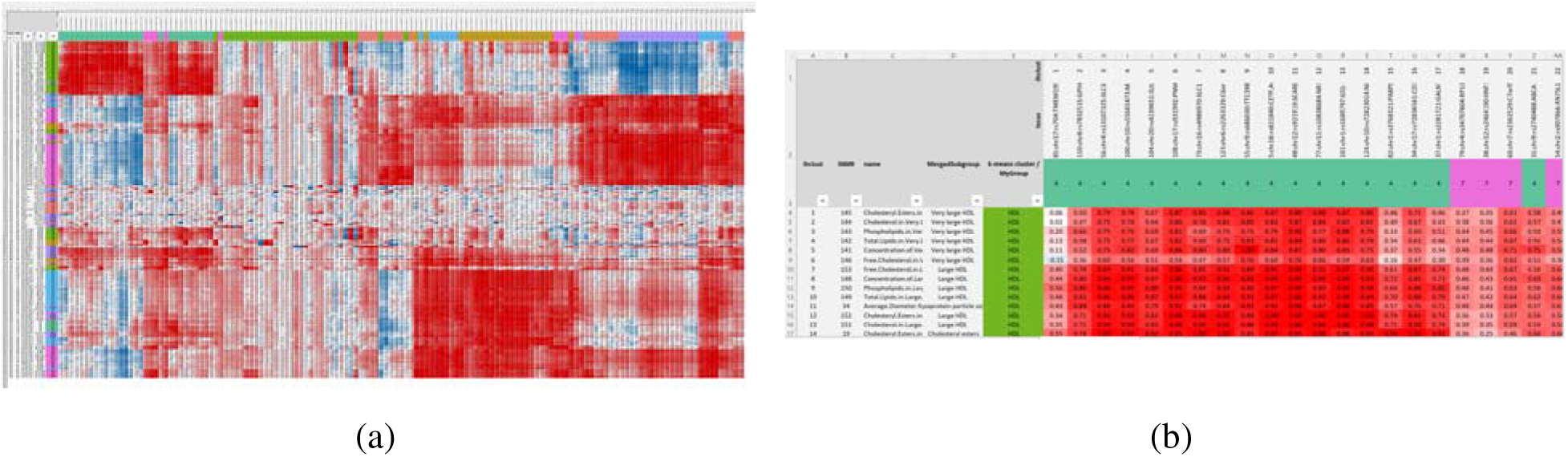
Association data for the 141 lead variants. The clustered and scaled effect sizes and -log10(p) values shown in Figure 1B**/C** are also available as data in Excel format (**Supplementary Tables 4&5**). The full association summary statistics for all NMR trait and ratio associations with all variants and all models reported in ST3 of Graham *et al*. are accessible as **Supplementary Data** on FigShare at https://doi.org/10.6084/m9.figshare.19728991.

**Supplementary Figure 4:**
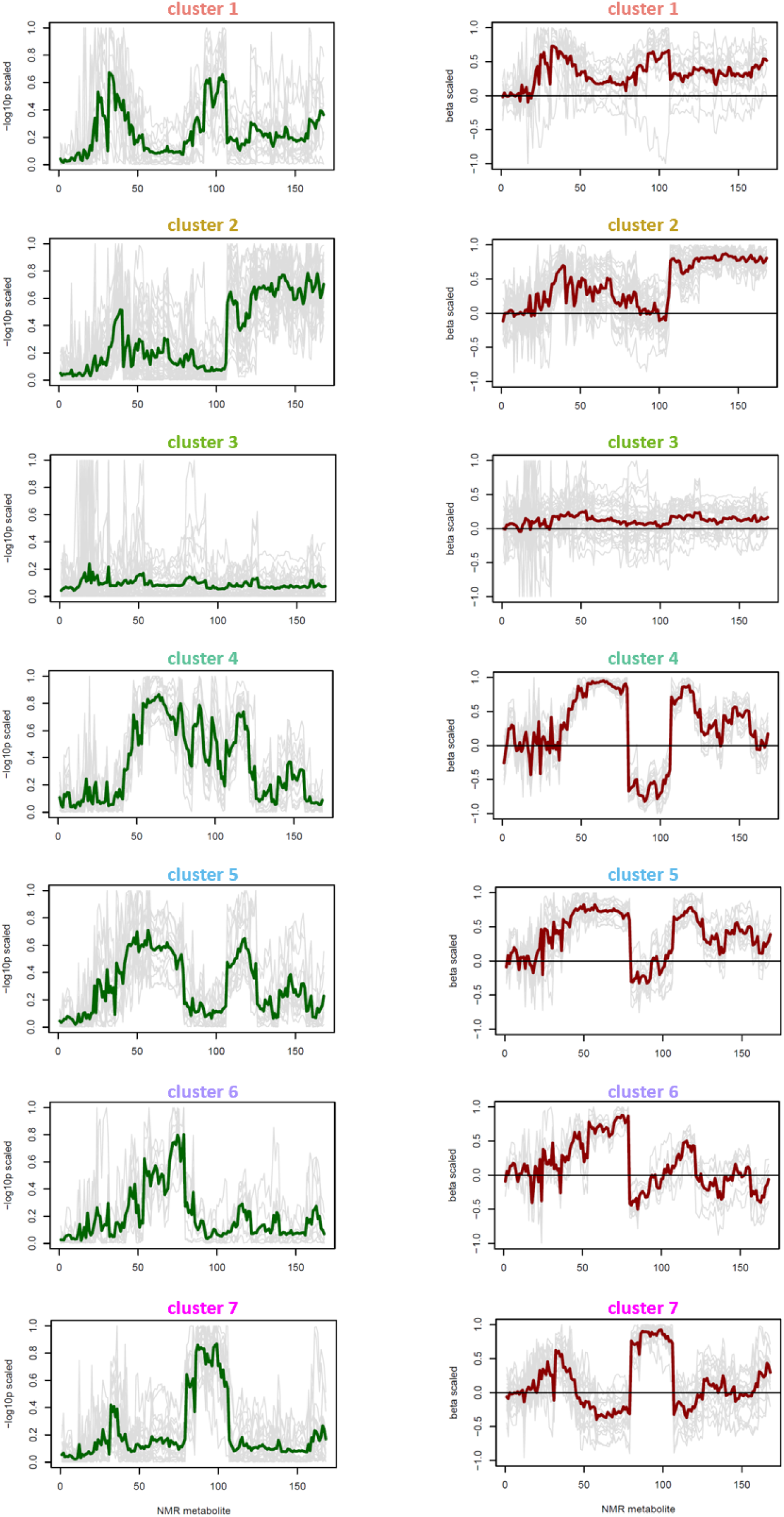
NMR association profiles by cluster. Individual association profiles are in grey and correspond to loci (columns) of the heatmaps presented in Figure 1B and 1C; NMR traits are ordered as in Figure 1 and **Supplementary Tables 4 and 5**; Average profiles for each cluster in green (normalized -log10(p-values)) and red (normal effect estimates, beta).

**Supplementary Figure 5:**
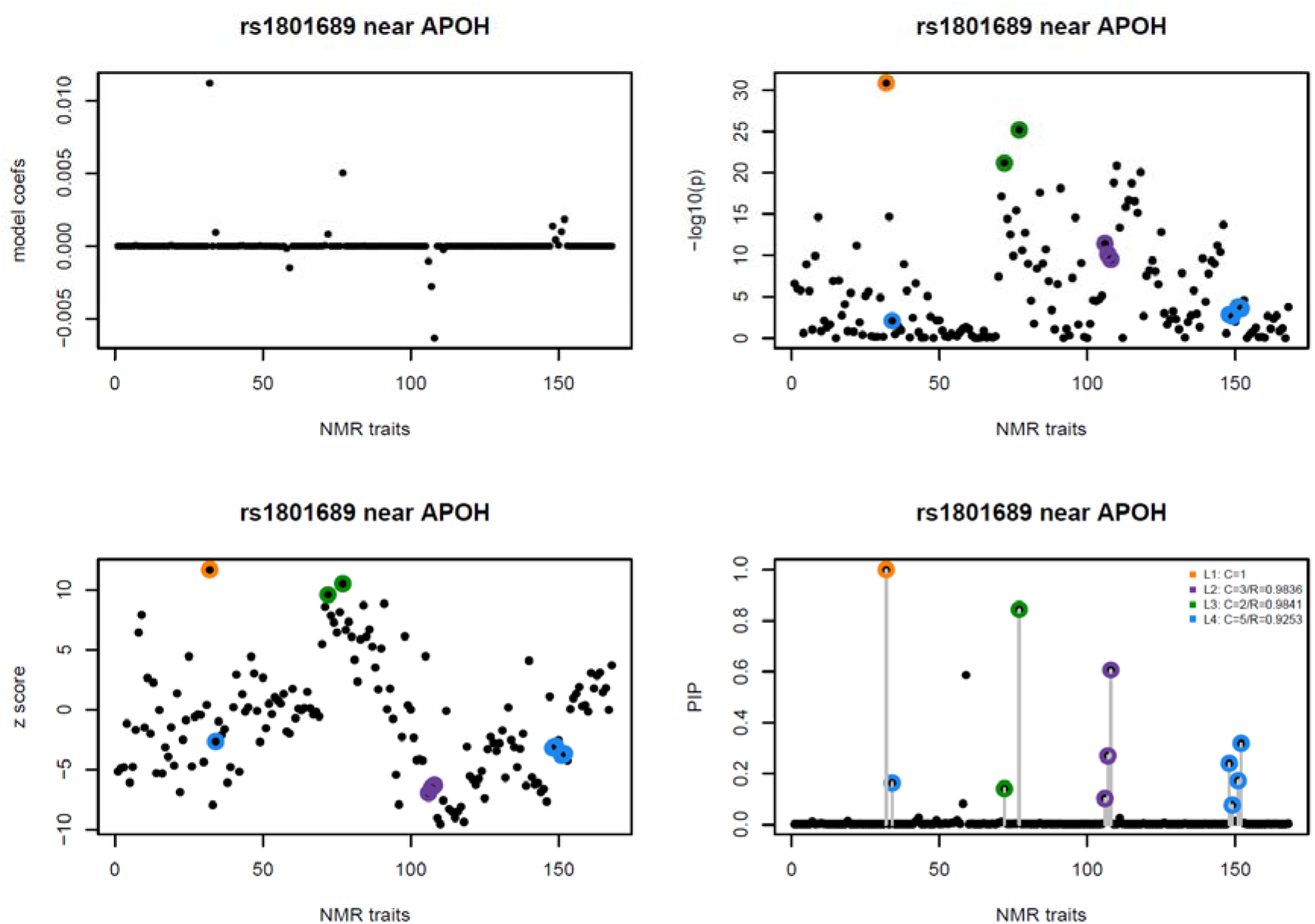
Example of a set of SuSiE analysis plots, showing model coefficients, p-values (-log10(p)) and z-scores for the single effect models, and the posterior inclusion probabilities (PIP) for the NMR traits (ordered on the x-axis as in **Supplementary Table 1**); Credible sets are colored and labeled; Details of credible set membership are in **Supplementary Table 6**; SuSiE analysis plots are provided in PDF format for all 141 lead associations on FigShare (file: run_susieR.pdf).

**Supplementary Figure 6:**
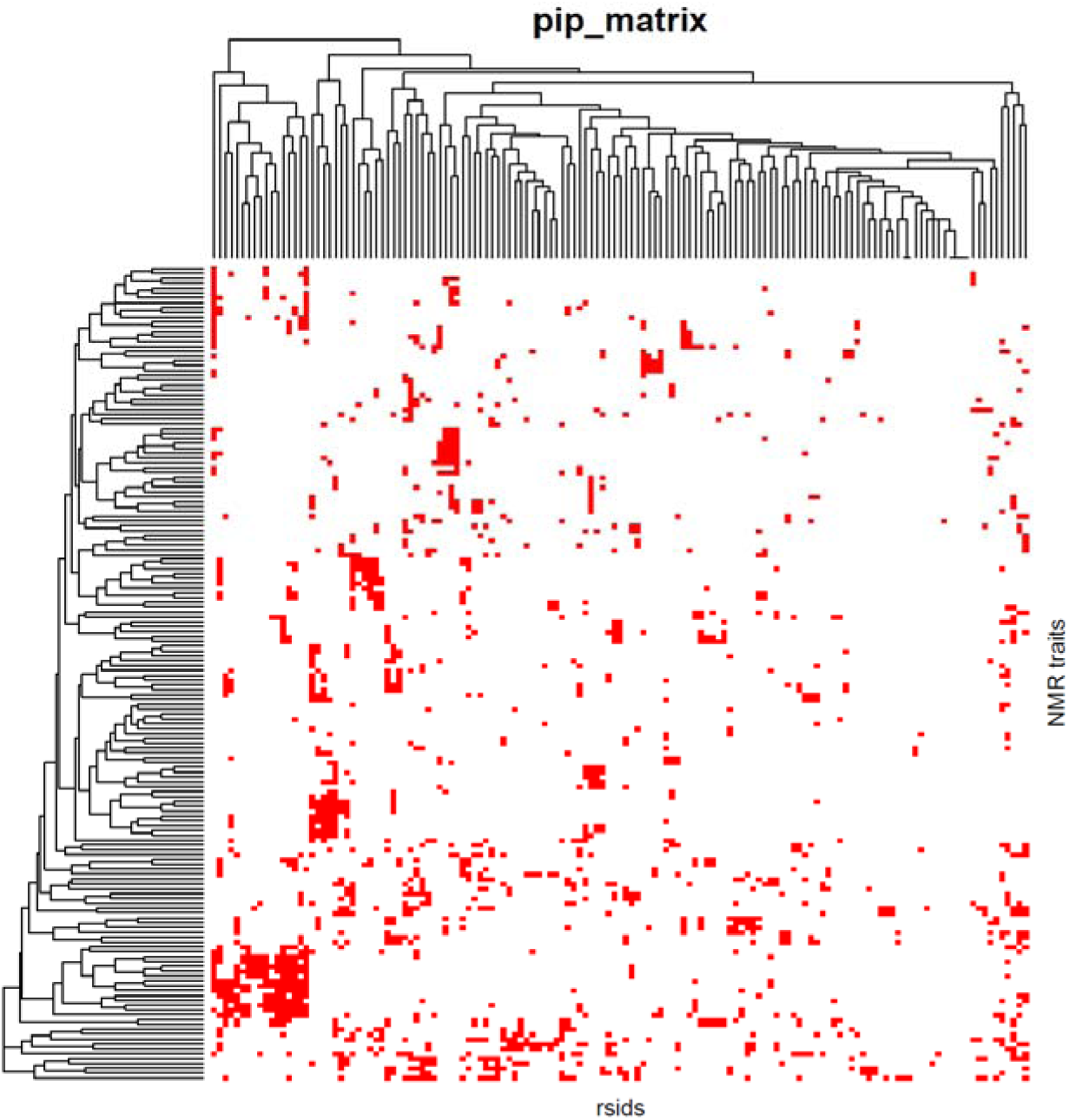
Binarized posterior inclusion probability (PIP) matrix for the SuSiE analysis of the 141 loci; red dots indicate traits that were included in the respective SuSiE models (**Supplementary Table 6**); The clustered matrix is also available as data in Excel format as **Supplementary Table 7**.

**Supplementary Figure 7.**
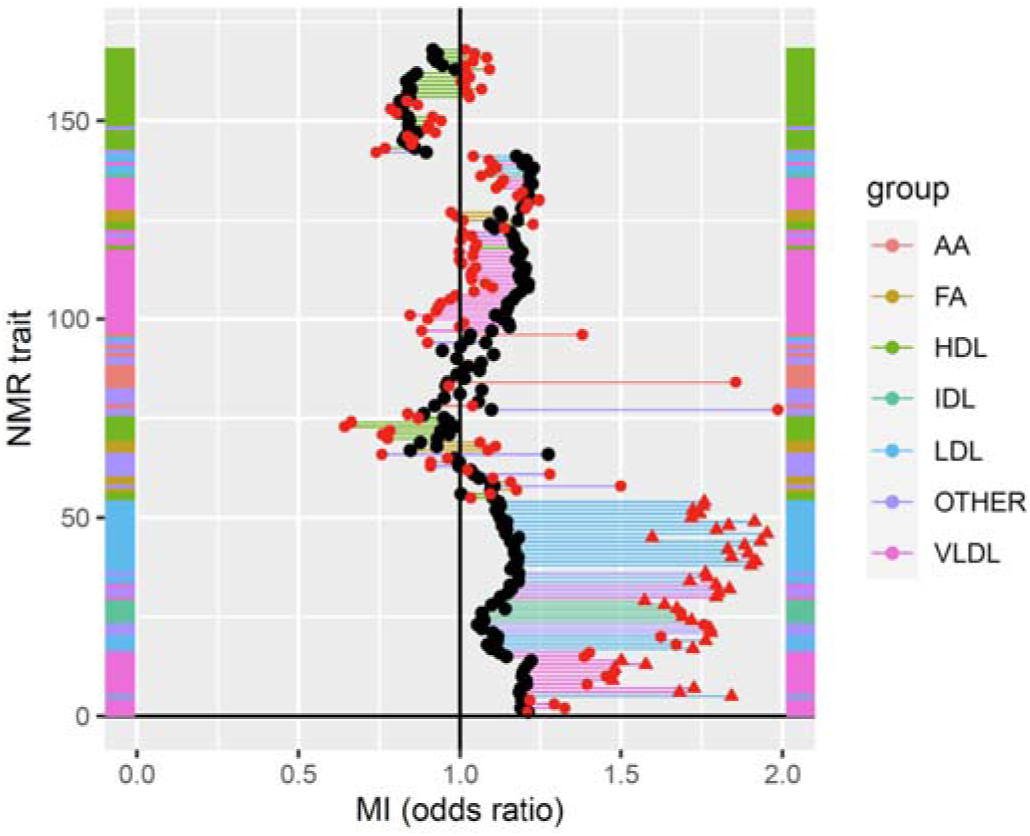
Mendelian randomization with MR. Odds ratios for the association of incident MI with the NMR traits (black) and estimates using Mendelian randomization (red); Triangles indicate Bonferroni significant MR estimates; Points for the same traits are connected by lines that are colored by the NMR trait group; Traits are ordered as in Figure 1B**/C**.

**Supplementary Figure 8.**
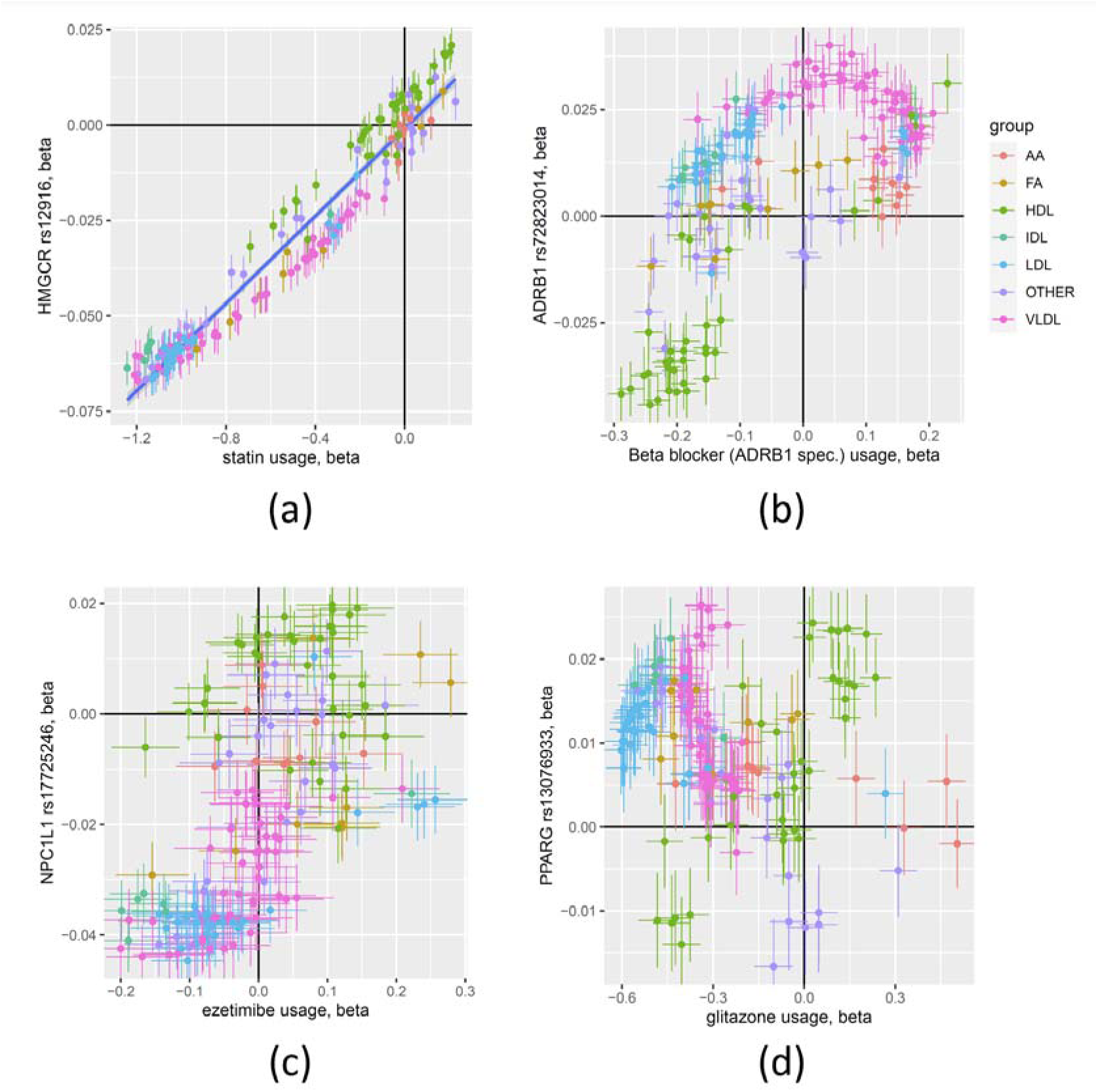
Association of variants in drug target genes and the corresponding drug usage. Scatterplots of the effect estimates for the associations of four variants near genes coding for established drug targets (HMGCR, ADRB1, NPC1L1, PPARG) with the 168 NMR traits and the effect sizes for the associations of the corresponding drugs (statin, beta blocker, ezetimibe, glitazone) with the 168 NMR traits (see legend to Figure 4); The statistical estimates are based on 15,050 statin users, 394 glitazone users (209 pioglitazone, 185 rosiglitazone), 615 ezetimibe users, and 5,939 users of ADRB1 specific beta blockers; These plots can be used to interpret how well a given variant possibly models the actual effect of the respective drug on the respective NMR traits.

**Supplementary Figure 9.**
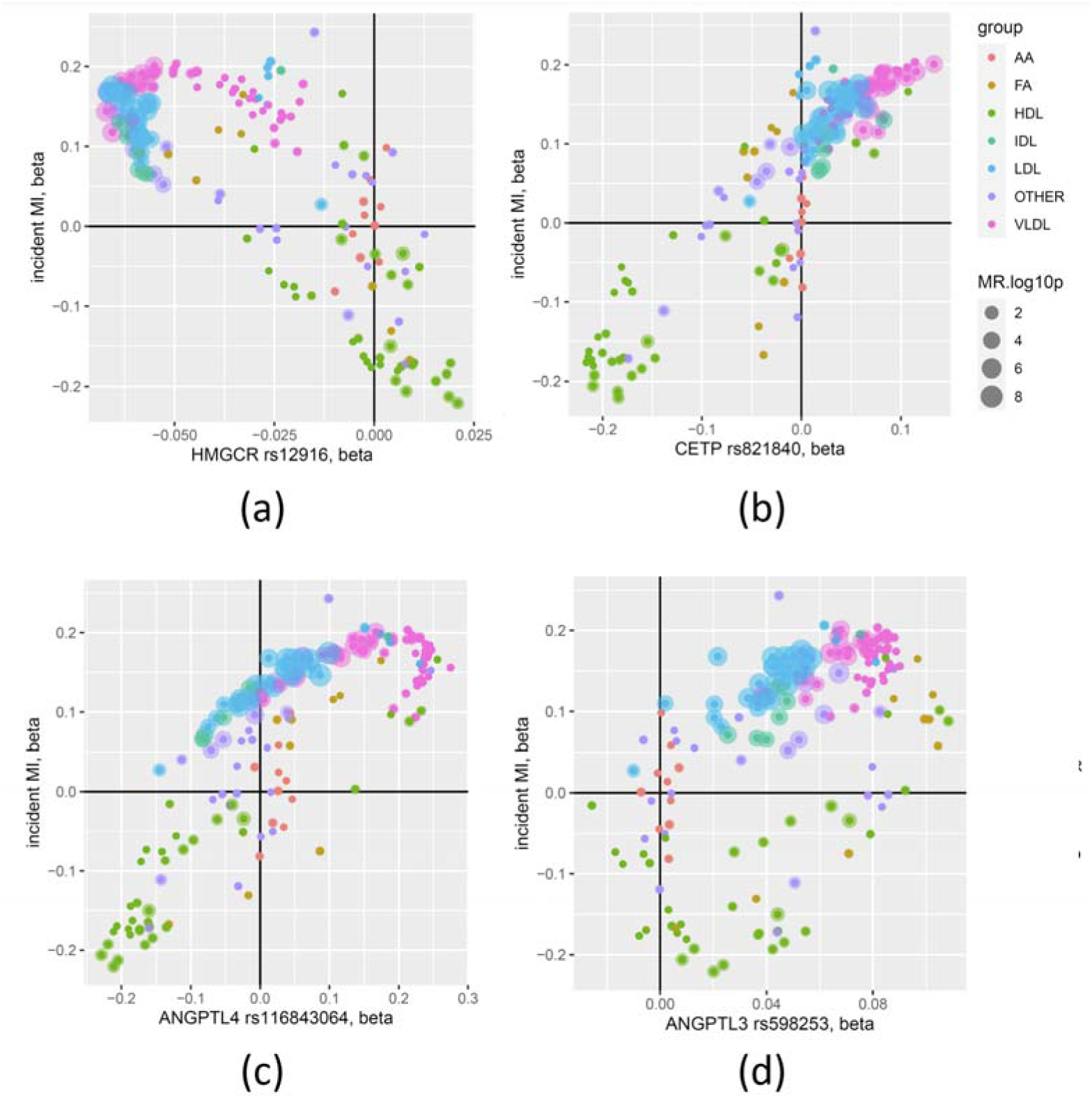
Association of variants in potential drug target genes and incident MI. Scatterplots of the effect estimates for the association of four variants near genes coding for established or potential drug targets (HMGCR, CETP, ANGPTL4, ANGPTL3) with the 168 NMR traits and the effect for the association of incident MI with the 168 NMR traits; Dot size represent the log10(p-value) of the MR estimates (MR Egger) for the MR with MI as an outcome; These plots can be used to interpret how the effect of genetic modulation of a potential drug target on the NMR traits aligns with the association of incident MR, and which of these traits are potentially causal, based the MR.

**Supplementary Figure 10.**
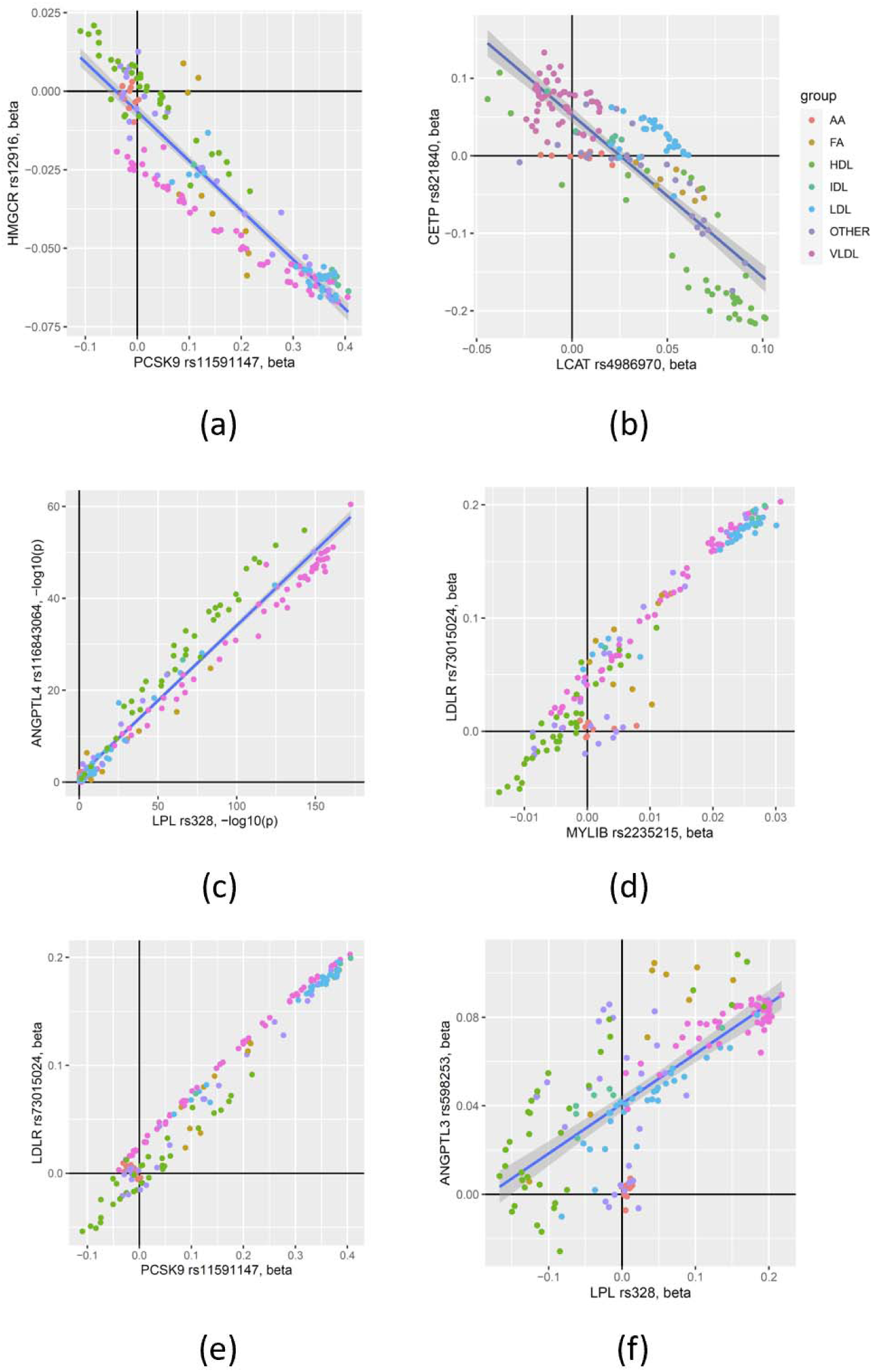
Scatterplots of the effect sizes for associations of selected pairs of variants with the 168 NMR-traits. These examples show the degrees of similarity that can be expected when targeting the respective gene products; They can be used to hypothesize on the potential effect of inhibiting a new target based on the profile(s) of known ones.

**Supplementary Figure 11.**
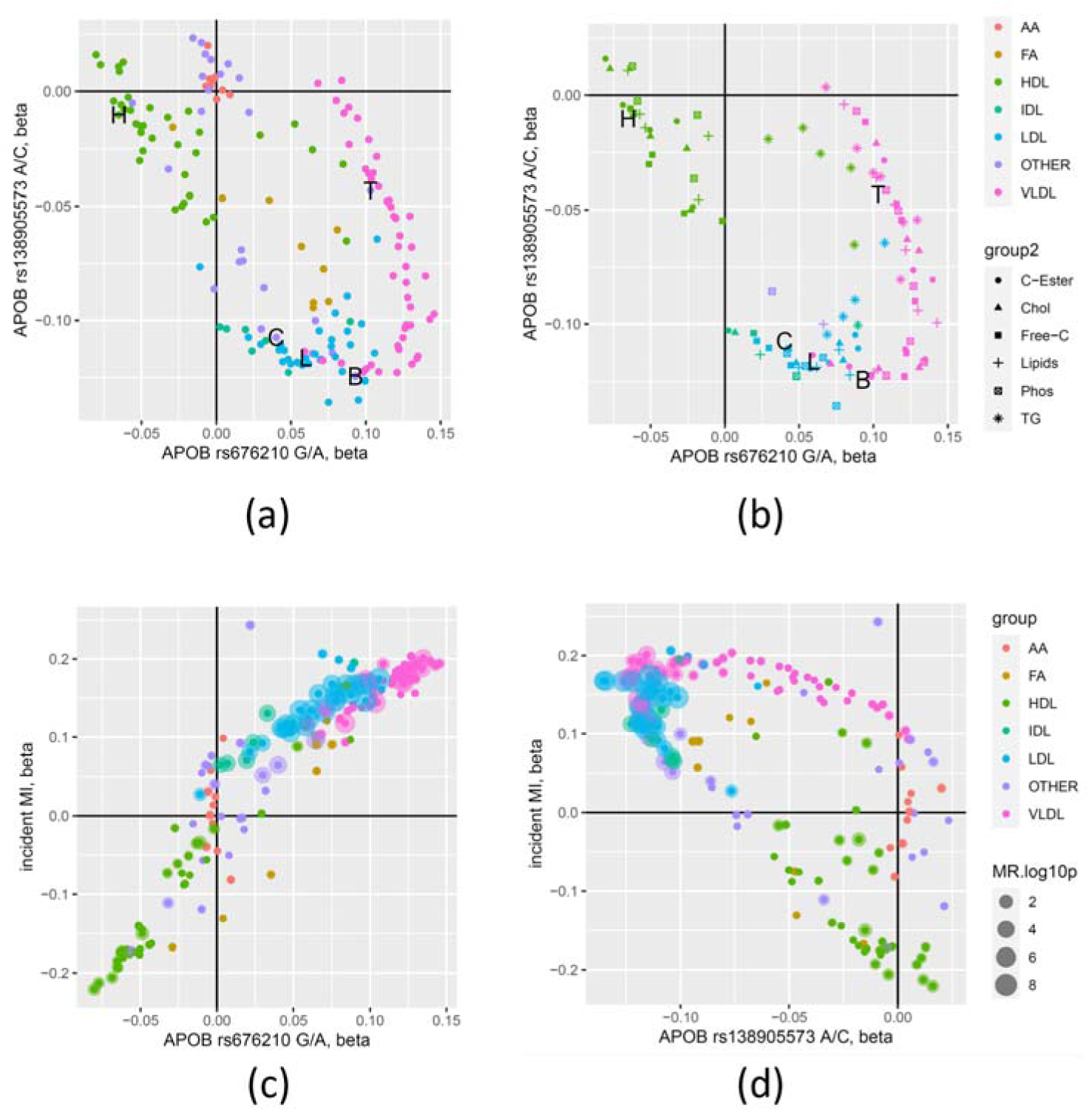
Association plots for the APOB locus. (a) Scatterplot of the effect estimates for the association of two genetically uncorrelated variants (rs676210 and rs138905573) at the APOB locus with the 168 NMR traits; The associations with the blood levels of HDL-C (H), LDL-C (L), total cholesterol (C), total triglycerides (T), and apolipoprotein B (B) measured by clinical biochemistry are indicated by letters; (b) as (a), but limited to lipid traits (symbols) in the respective lipoprotein particles; (c,d) Scatterplot of the effect estimates of the respective APOB variants and the 168 NMR-traits and the association of incident MI with the NMR-traits; Dot sizes represent the log10(p-value) of the MR estimates (MR Egger) for the MR with MI as outcome; Note that rs676210 is a protein altering variant in the APOB gene while rs138905573 is located upstream of APOB; This example shows that two variants located to a same gene locus can have very different effects; In this case the protein altering variant may for instance change the physical binding properties of APOB and its interaction with other molecules in the lipoprotein particles, while the non-coding variant may alter APOB protein levels altogether.

**Supplementary Figure 12:**
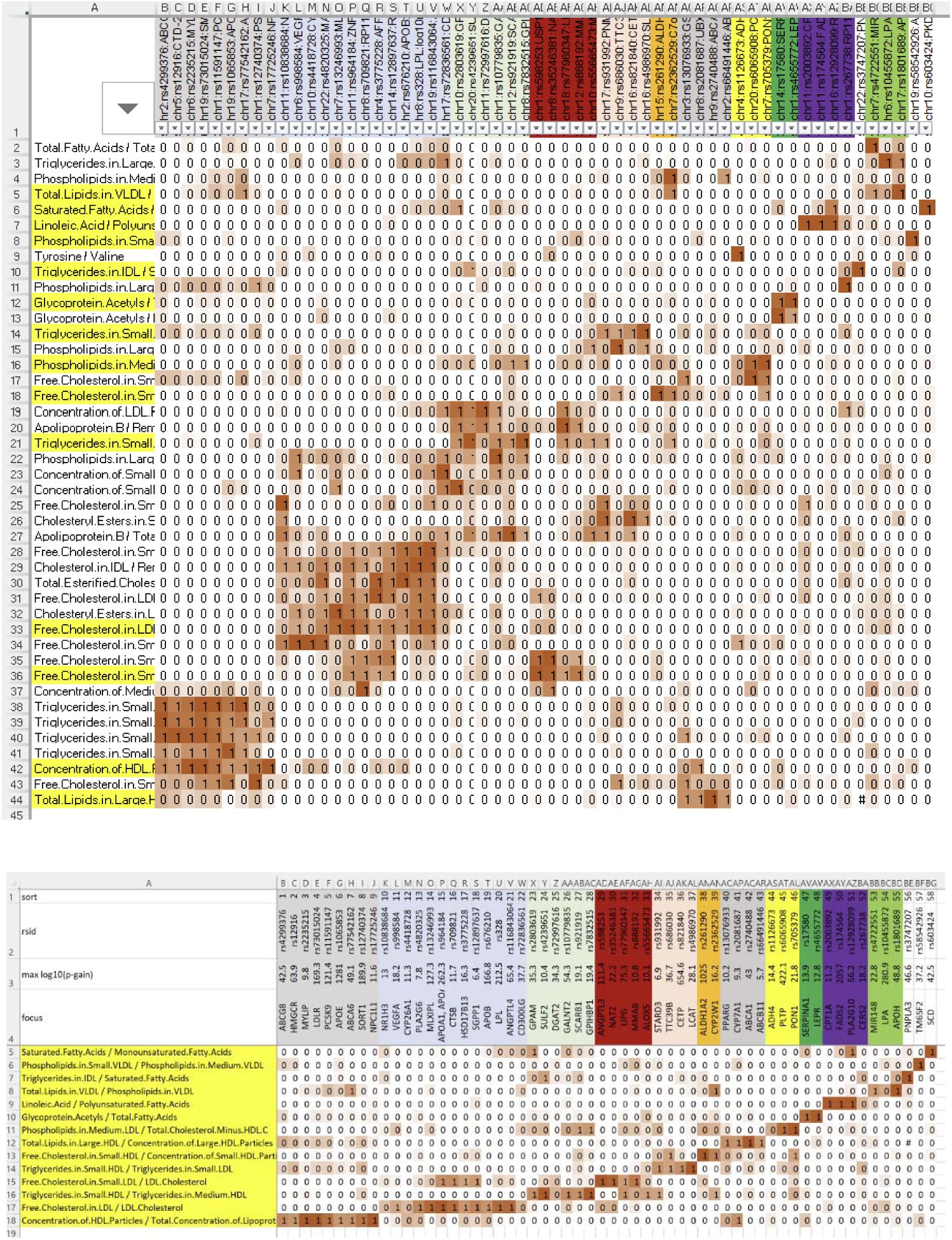
Identification of the 14 key ratios. (yellow) that are associated with the 58 genes (clusters highlighted as colored columns) involved in lipoprotein metabolism, transport, and remodeling; full matrix with all ratios (top) and limited to the ratios with the largest p-gain per cluster (bottom); matrix entries are normalized log10(p-gain) values; The data is also available in Excel format as **Supplementary Tables 11&12**.

**Supplementary Figure 13.**
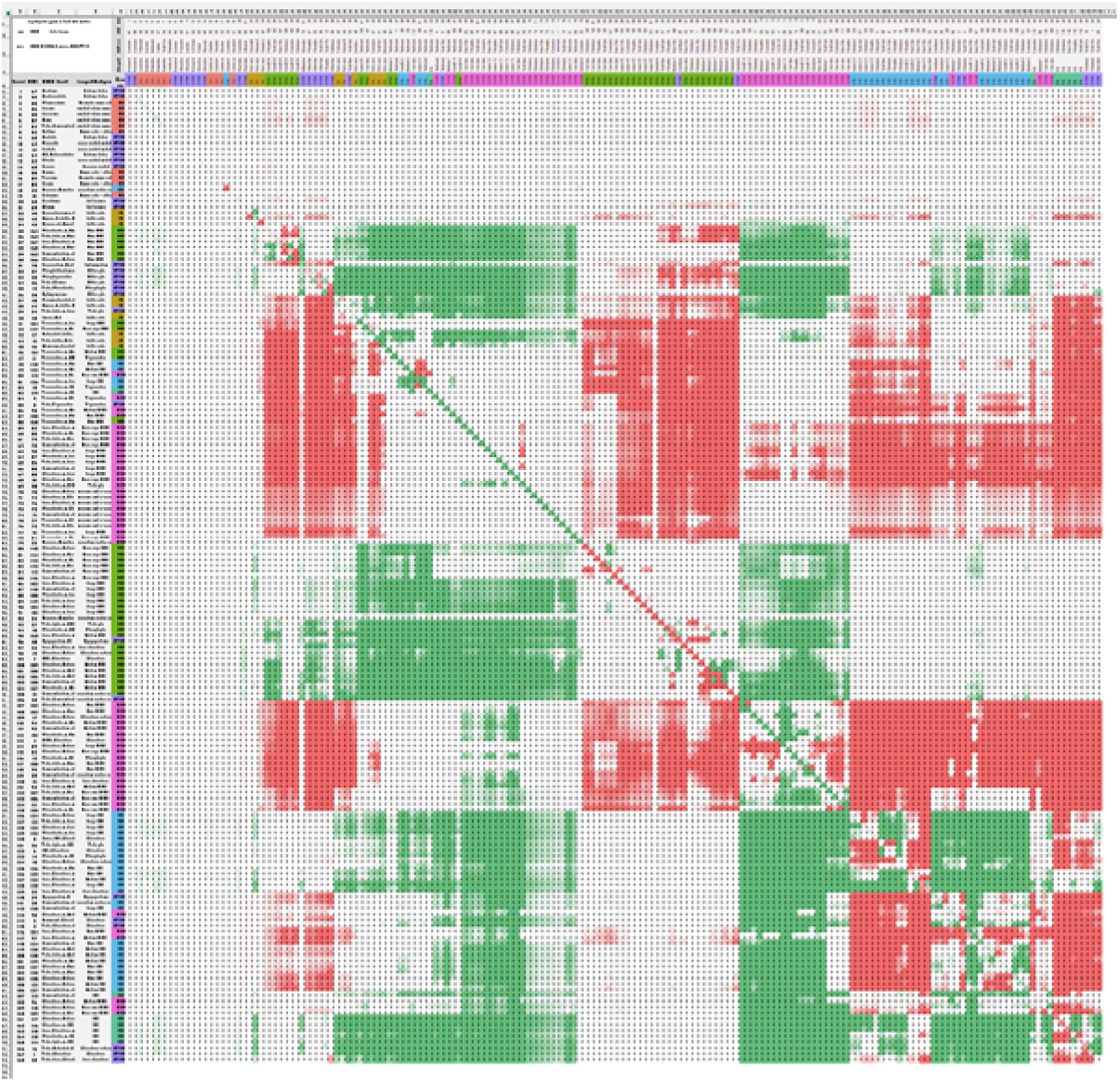
P-gain matrix. An example of a 168×168 matrix containing all log10(p-gain) values for the association of one variant (rs116843064 near ANGPTL4) with all 14,196 traits and ratios; The diagonal contains the -log10(p-values) and the off-diagonal cells contain the log10(p-gain) values, where the sign indicates the direction of the association; Formatted data for this example is in **Supplementary Table 15**; Data for all loci is available on Figshare at https://doi.org/10.6084/m9.figshare.19728991.

**Supplementary Figure 14:**
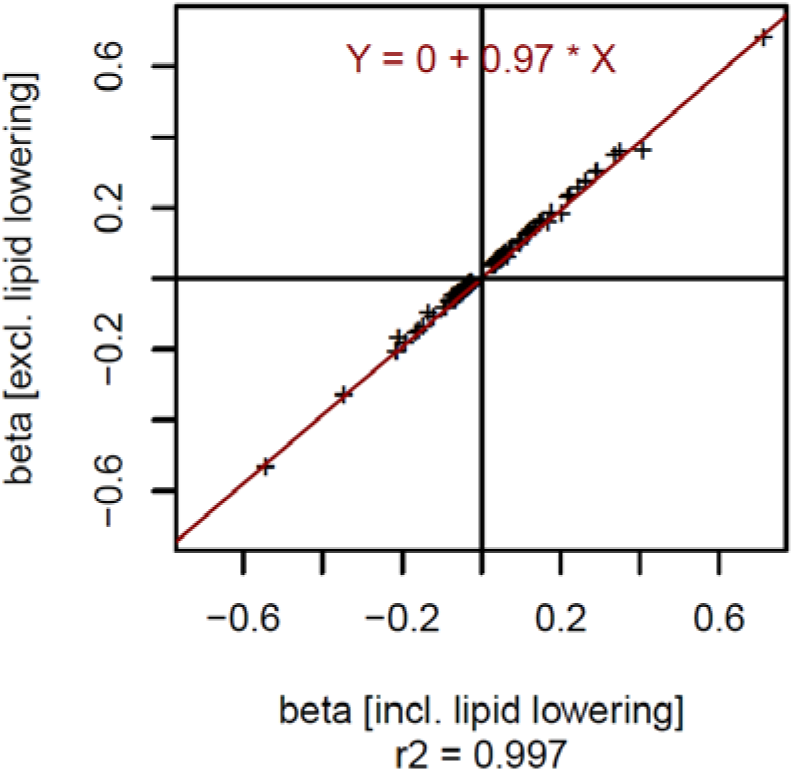
Effect of lipid lowering medication as a covariate. Scatterplot of the effect sizes (beta) for the associations with the respective lead NMR trait at the 141 loci, including and excluding lipid lowering medication as a covariate; This plot shows that major biases introduced by including lipid lowering medication as a covariate can be ruled out.

## SUPPLEMENTARY TABLES

Supplementary tables are provided in EXCEL format.

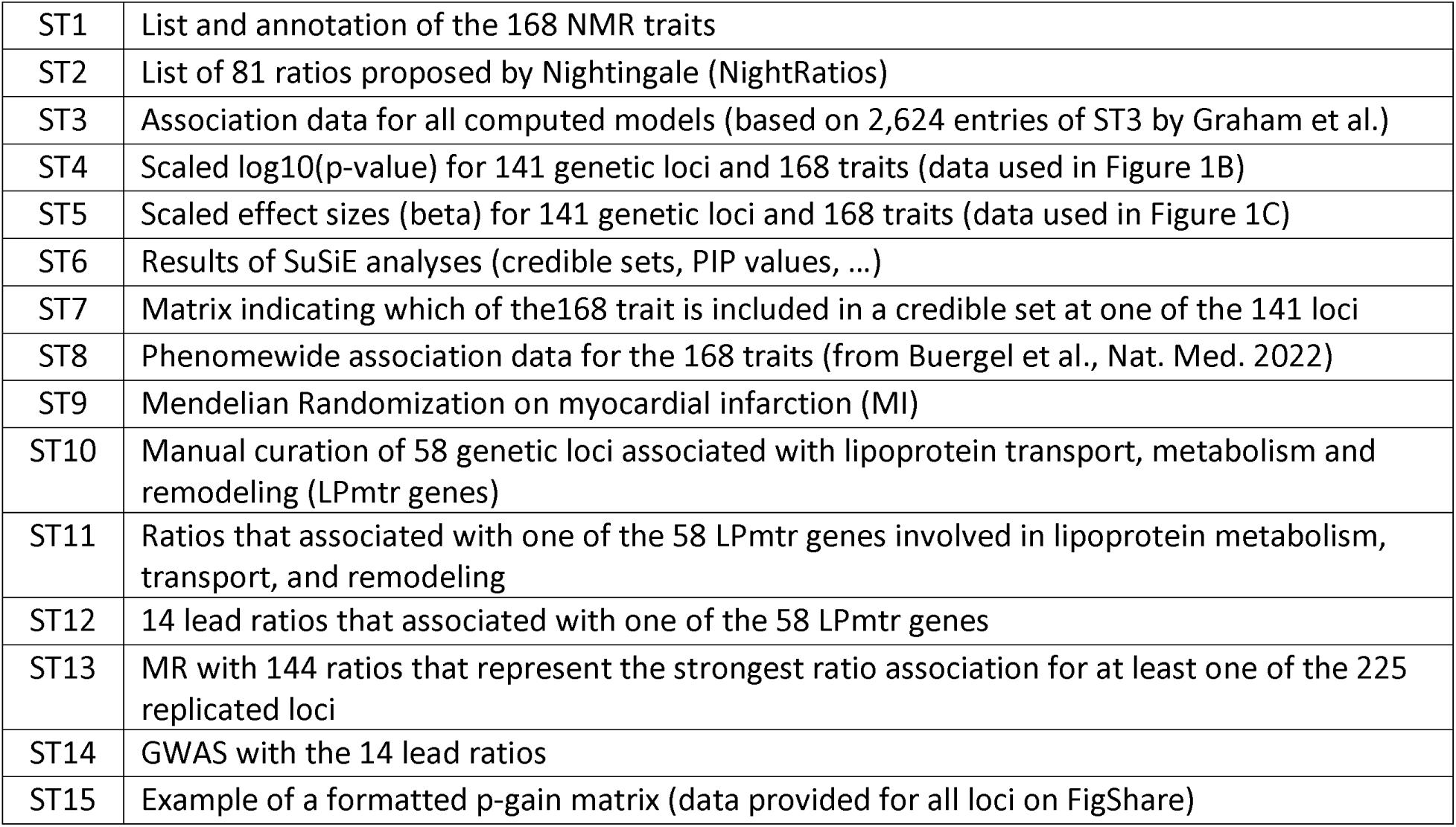

## SUPPLEMENTARY DATA

Summary statistics for all NMR-trait and NMR-ratio associations at all 1,906 loci, ratio association matrices similar to **Supplementary Table 15** in tab-separated text format, as well as all MR instruments in MRBase format are available on FigShare https://doi.org/10.6084/m9.figshare.19728991.

